# Risk of plague transmission from human cadavers: a systematic review

**DOI:** 10.1101/2021.02.21.21252152

**Authors:** Sophie Jullien, Nipun Lakshitha de Silva, Paul Garner

## Abstract

Plague can be fatal. Ebola has raised concern of the risk of infection from bodies of people that have died from other dangerous infectious diseases. We sought to estimate the risk of human remains from someone who was infected with plague transmitting *Yersinia pestis*. With no clear direct evidence, we developed a causal chain and carefully searched and assessed the literature at each step: we assessed the infectiousness of body fluids of people ill with plague; we sought for reported infection acquired from human and animal cadavers; and examined evidence of body fluid infectiousness of cadavers, seeking any information about the length of infectiousness. We concluded that pneumonic plague can occur after intense manipulation of the cadaver, presumably from inhalation of respiratory droplets; and that bubonic plague can occur after contact with infected blood in people with skin cuts or abrasions. Establishing a quantitative measure of risk was not possible from the evidence available.

**Author summary:** Plague is an infectious disease that is of particular concern due to its high risk for human outbreaks. Understanding the level and duration of infectiousness of the body of someone who was infected with plague is essential to establish adequate preventive measures while handling corpses. We found limited evidence that human cadavers who have died from plague remain infectious, both from inhalation of particles generated by handling and by direct skin contact.

## Introduction

Plague is an ancient disease that has killed millions in the past [1]. Plague remains a current threat in many parts of the world [2], and was categorized by the World Health Organization (WHO) as a re-emerging disease [3]. Caused by *Yersinia pestis*, a non-motile gram-negative coccobacillus, the disease is a zoonosis with rodents as the main reservoir [4,5], and is transmitted to human by infected rodent fleas, although it can also be transmitted by bites from infected animals and by handling, ingestion, and inhalation of aerosolized droplets from infected tissues, animals or humans (Fig 1) [6–10].

**Fig 1.**
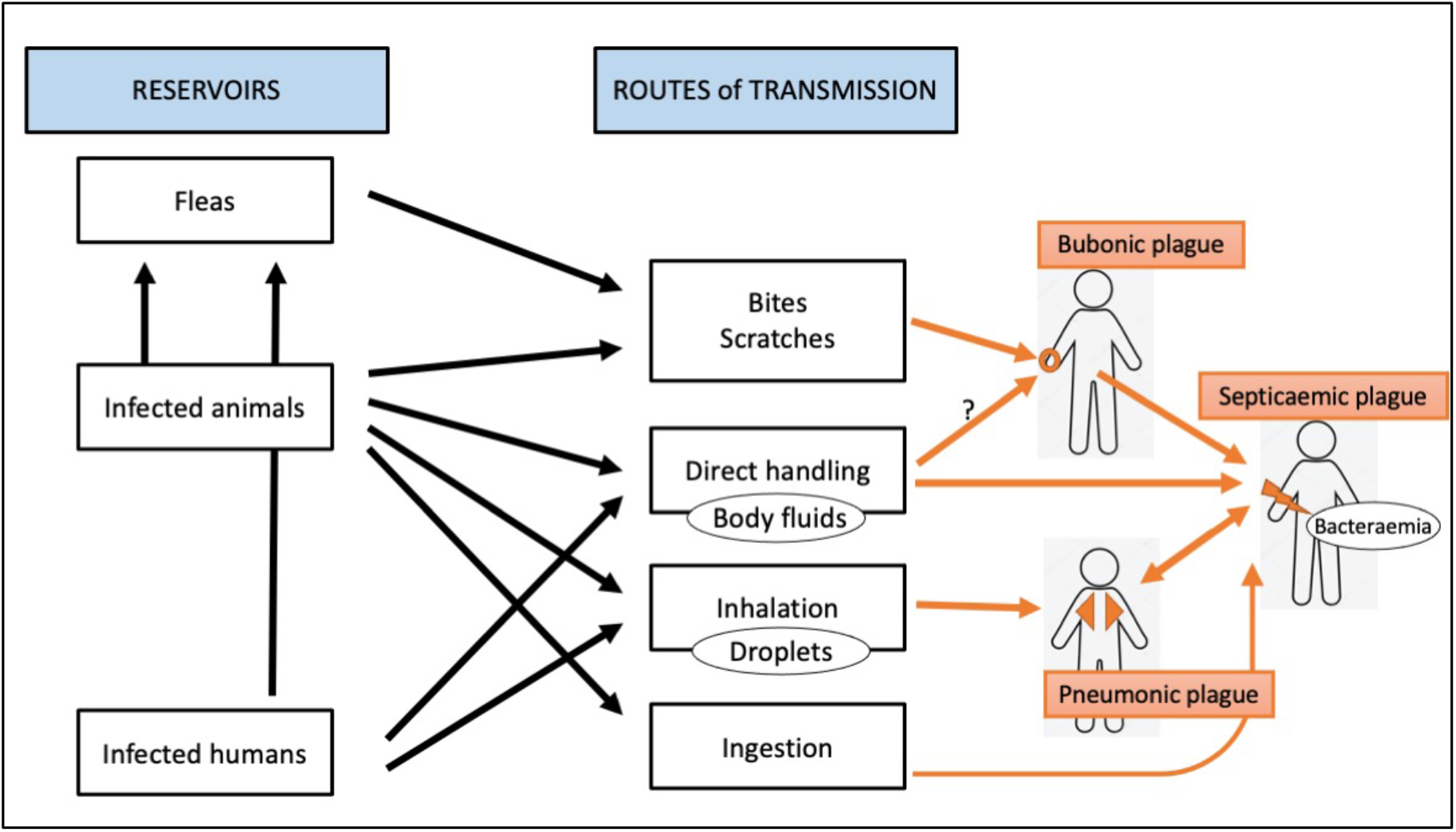
Reservoirs of *Y. pestis* and routes of transmissions leading to the different forms of plague.

Plague may present as bubonic plague, with lymph nodes inflammation following a flea bite or scratch from an infected animal [11,12]; pneumonic plague from inhalation of droplets from infected humans or animals; or septicaemic plague, from the hematogenous spread of bubonic or septicaemic plague [13]. The question as to whether dead bodies are infectious has become current, with the Ebola outbreak heightening concerns. Very little is known about how contagious human cadavers are and for how long, and by which way transmission can occur - and often relatives may use no protection in traditional funeral rituals. Plague transmission from an infected human cadaver can possibly occur through three routes: contact with infectious body fluids directly (through skin or inhalation) or indirectly (through contaminated clothing), or bites from infected fleas from the cadavers (on the body or in the clothes). In this review, we aimed to estimate the risk of plague-infected human cadavers transmitting *Y. pestis* through infectious body fluids to help a WHO Guideline Group make recommendations for protective personal equipment for health workers. As we anticipated that direct evidence from human cadavers is lacking, we developed the approach through a logic framework (Fig 2) and a full protocol prepared in advance (registered in Prospero [14]), with three review parts: infectiousness of different body fluids of people ill with plague (part 1); reported infection acquired from human and animal cadavers (part 2); and evidence of body fluid infectiousness of animal or human cadavers, including length of cadaver infectiousness (part 3).

**Fig 2.**
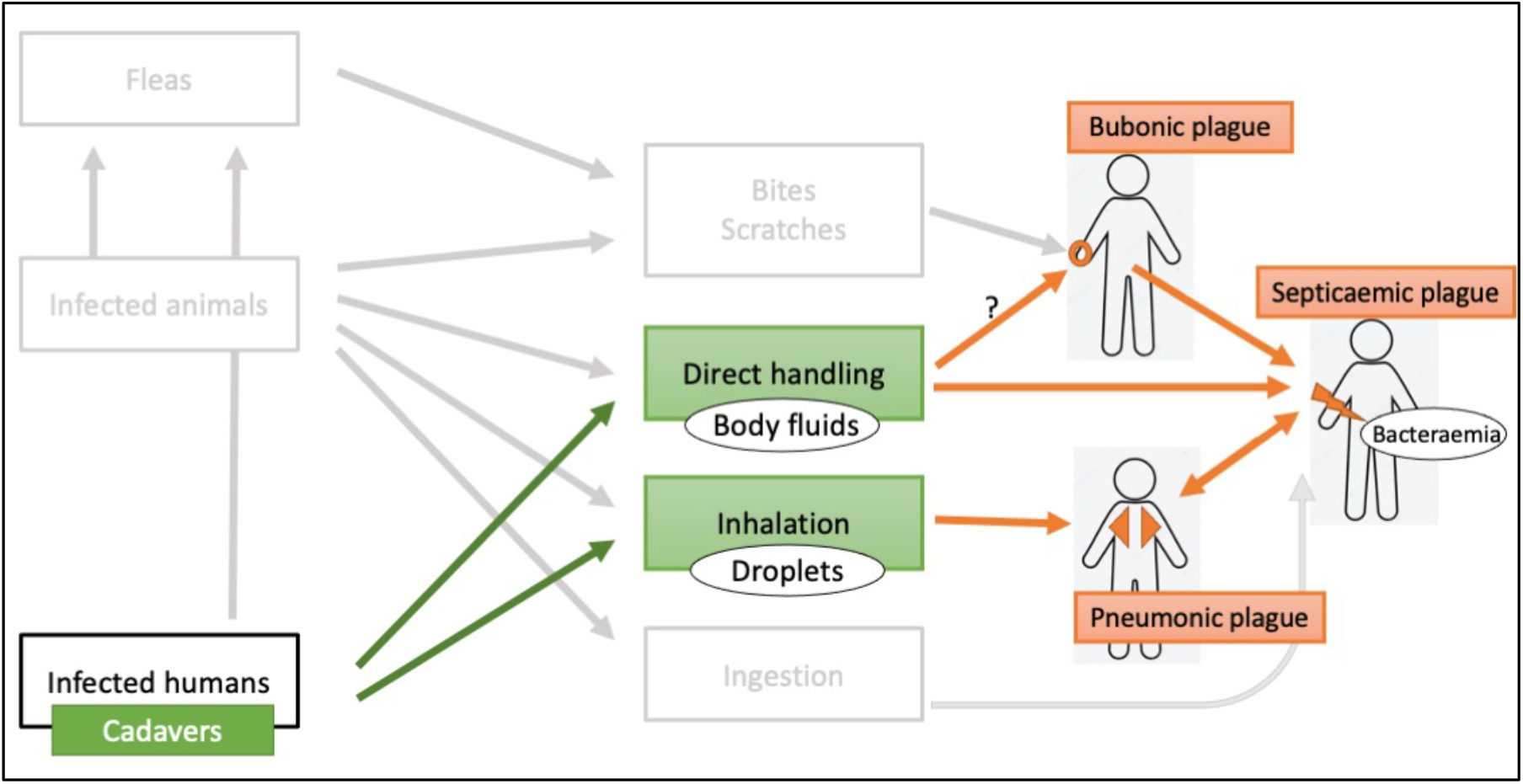
Potential transmission routes from human cadavers.

## Methods

For each of the three parts, we developed inclusion criteria (Table 1). We excluded cases where plague infection was clearly attributed to consumption of infected meat, as we estimated that the risk of infection acquired from consumption of human cadavers was unlikely. We also excluded cases of vector transmission such as fleas.

**Table 1.** Inclusion criteria for the three parts of the review

|  | Part 1. Infectiousness of body fluids of people ill with plague | Part 2. Infection acquired from human and animal cadavers | Part 3. Body fluid infectiousness of animal or human cadavers, including length of cadaver infectiousness |
| --- | --- | --- | --- |
| **Types of study** | Descriptive studies including surveillance data, case series and case reports | Descriptive studies including case series and case reports | Descriptive studies including case series and case reports |
| **Participants** | People with confirmed plague | People or animal that have died from confirmed plague | People or animal that have died from confirmed plague |
| **Outcomes** | New case of human with confirmed plague attributed to direct transmission from infected human (human-to human transmission) | New case of human with confirmed plague attributed to direct transmission from remains of a person or animal that was infected with plague | - New case of human with confirmed plague attributed to direct transmission from remains of a person or animal that was infected with plague, where time period between the time of death of the plague victim and time of contact through which transmission occurred is specified.<br>- Isolation of *Y. pestis* by culture from body fluids from remains of a person or animal that was infected with plague, where time period between the time of death of the plague victim and the time of identification of *Y. pestis* in the body fluid is specified. |
| **Exclusion criteria** | None | Studies reporting only cases of plague attributed to consumption of infected meat, or cases of vector transmission such as fleas | Studies looking at persistence of *Y. pestis* DNA in old cadavers that were already buried as well as persistence of *Y. pestis* in the soil or environmental surfaces |

### Search methods for identification of studies

We conducted the search up to the 20 May 2019 and identified all relevant studies regardless of language, publication status, or publication date limit.

#### Electronic searches

We searched the following databases using the search terms and strategy described in S1 Appendix: MEDLINE (PubMed), Embase (accessed via Ovid), Science Citation Index (Web of Science), and Scopus.

#### Searching other resources

We also hand searched the reference lists of all eligible papers and contacted relevant researchers working in the field.

### Data collection and analysis

#### Selection of studies

Two review authors (SJ and NS) independently screened all the abstracts retrieved by the search strategy using predefined eligibility criteria and allocated studies to the inclusion criteria they meet, which might include more than one of the three review parts. We retrieved full-text copies for studies remaining after the first screening and applied the predefined inclusion criteria. Manuscripts in French, Russian, German and Chinese were assessed by one of the review authors or plague experts for whom mother tongue was one of these languages, or with the help of online translation. We resolved any disagreements in assessment through discussion. We listed all studies excluded after full-text assessment in the “Characteristics of excluded studies” table (S2 Appendix). We illustrated the study selection process in a PRISMA diagram (Figure 3).

**Fig 3.**
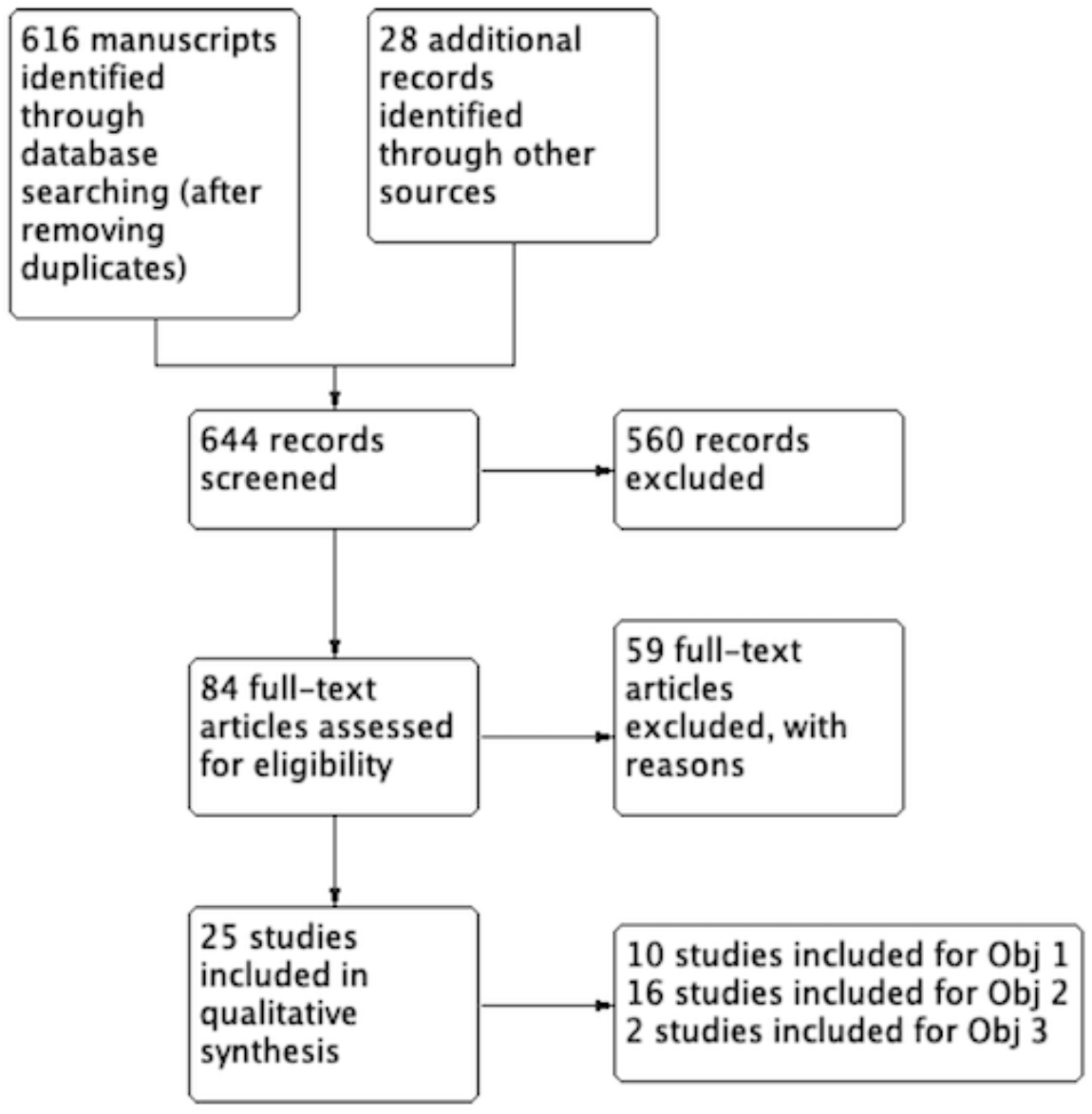
Study flow diagram.

#### Data extraction and management

Two review authors (SJ and NS) extracted data with data extraction forms. For each included study, we gathered information into “Characteristics of included studies” tables, comprising different data depending on the review part being addressed.

For studies in part one, we extracted data on setting, date, description of the index case, route of transmission mentioned by the study authors, methods used for contact tracing, contact definition, number of contacts identified and number of those infected, and any other relevant notes. For studies in part two, we extracted data on setting, date, source of infection, duration of exposure, route of transmission mentioned by the study authors, time from death of the infective human or animal cadaver to moment of exposure, basic characteristics of cases infected, and any other relevant notes. For studies in part three, we extracted data on setting, date, type of body fluid contaminated, duration of persistence of *Y. pestis* in body fluids between death and contact or *Y. pestis* identification, method of diagnosis of plague in the cadaver or of identification of *Y. pestis* from body fluids, route of transmission mentioned by the study authors, and any other relevant notes.

#### Assessment of risk of bias

Two review authors (SJ and NS) assessed aspects of risk of bias of each included study, to consider the study limitations when making conclusions about the results. We used a simple appraisal tool comprising of six questions, modified from the quality appraisal tool developed by Cho et al that has been used for assessing risk of bias of descriptive studies [15] (S3 Appendix).

#### Data synthesis

No statistical analysis was possible, and data were summarized narratively in text and tables for each of the three parts.

## Results

### Result of the search

We identified 644 studies (616 records from the literature search after removing duplicates and 28 additional studies by hand search) and included 25 in the review (Fig 3). Ten of them addressed the part one, 16 of them addressed the part two and two of them addressed the part three, with three studies addressing more than one part.

### 1) Infectiousness of body fluids of people ill with plague

#### Study description

Ten studies report cases of direct human to human transmission of plague demonstrating infectiousness of body fluids of people ill with plague (details in S4 and S5 Appendices). There are four studies describing plague cases from outbreaks or surveillance registries from the 20^th^ century in Brazil [16], South Africa [17] and the Unites States [18,19] and six reporting more recent outbreaks between 1997 and 2017 in Madagascar [20– 23], Uganda [24], and the Democratic Republic of the Congo [25]. Overall, the ten studies described a total of 2388 humans who became infected by plague attributed to direct contact with humans, including an outbreak in Madagascar with 1861 cases [20]. All the infected cases had primary pneumonic plague, except for four cases with septicaemic plague [17,25]; and six cases with mixed forms described with probable pneumonic affectation secondary to buboes [17].

#### Risk of bias

Six studies had adequate description of patient characteristics such as age, gender and forms of plague, three had incomplete description, and one study did not provide such information (Table 2, details in S5 Appendix). In four studies, there were efforts to trace contacts from the index case, suggesting a certain level of contagiousness of the disease by assessing infected and non-infected people from the same index cases. Laboratory methods for confirming infected cases of plague were part of our inclusion criteria, although only partially detailed in two studies. We judged that the route of transmission and that the causality of it for plague infection was plausible in eight studies. In two studies (including 50 cases), this was difficult to judge based on the limited information provided.

**Table 2.** Risk of bias summary for studies on human to human transmission of plague (part 1)

| Study ID | Were patient characteristics adequately reported? | Was there some effort to trace all contacts from the index case? | Were the methods used for tracing contacts adequate? | Were the laboratory methods used for defining a confirmed case of plague reliable? | Was the route of transmission plausible? | Was the cause-effect of transmission plausible? |
| --- | --- | --- | --- | --- | --- | --- |
| Almeida 1981 | Partial | Unknown | Not applicable | Yes | No | Unknown |
| Begier 2006 | Yes | Yes | Yes | Partial | Yes | Yes |
| Bertherat 2011 | No | Partial | Unknown | Yes | Yes | Yes |
| Evans 2018 | Yes | Unknown | Not applicable | Yes | Yes | Yes |
| Kellogg 1920 | Yes | Unknown | Not applicable | Yes | Yes | Yes |
| Kugeler 2015 | Partial | Unknown | Not applicable | Yes | Unknown | Unknown |
| Rabaan 2019 | Partial | Partial | Unknown | Yes | Yes | Yes |
| Ramasindrazana 2017 | Yes | Yes | Yes | Yes | Yes | Yes |
| Ratsitorahina 2000 | Yes | Yes | Unknown | Partial | Yes | Yes |
| Richard 2015 | Yes | Yes | Unknown | Yes | Yes | Yes |

#### Findings

Bloody sputum was described from the index case [22,24], from infected contacts [17,21], and from both [23]. The study authors attributed the route of transmission from the index cases to the infected contacts to ‘respiratory droplets’ or ‘direct transmission through infective cough droplets’ in four studies [19,20,22,24] or to ‘aerosolized bacteria spread through coughing’ or ‘aerosol by droplets or by contaminated dust’ in two studies [23,25]. Two studies concluded that the infected cases were consistent with ‘human to human transmission’ without further details [18,21], and one study did not provide any suggestion for the route of transmission of plague [16]. There were not enough details provided by the study authors to understand whether the six mixed cases and the four cases of septicaemic plague were attributed to human to human transmission [17,25].

To assess how contagious are people sick with plague, we extracted data about people in contact with plague cases but who did not get infected. Across four studies that provided such information, 51 contacts got infected from five index cases (although some infected contacts acted as index cases for additional infected cases), and 341 contacts who were exposed to the five index cases did not develop plague [21–24]. The attack rate estimated by the study authors were 8%, 8.4% and 55% [22–24]. Transmission rate was assessed in a single study and reported to be 0.41 susceptible people per day [21]. While contacts did not receive chemoprophylaxis in one study [24], post-exposure prophylaxis was given to all contacts in one outbreak [22], to 39/41 contacts in another outbreak [23], and to 35 contacts with positive serology in a third outbreak [21]. When described, infected contacts had been in close and prolonged exposure to index cases, such as close family members, main care takers including nurses and physicians, or other family members or villagers staying at the same house as the plague cases [17,19,22–24]. Four studies from South Africa and Madagascar attributed plague transmission to activities related to funerals, such as ‘preparing bodies for funerals’ or ‘active participation in the funerals’ [17,20– 22]. Evans et al added that ‘the disease is transmitted to relatives, friends, or caregivers but not to more loosely associated contacts’ [17].

Contacts exposed to plague but who were not infected included family members who slept in the same bed as the plague patient until the night before their death [23,24] including persons who slept with their heads at a distance of less than two meters from the coughing plague patient [24].

#### Summary

The evidence shows that direct transmission through infective cough droplets from people ill with plague may occur, but sometimes this is only after close and prolonged exposure. We found no publication describing interhuman transmission of plague through other body fluids such as blood (although respiratory droplets from pneumonic cases come from grossly bloody sputum), urine, faeces, sweat or bubo pus.

### 2) Plague infection acquired from human and animal cadavers

#### Description of studies

We included 16 studies (details in S6 and S7 Appendices), all retrospective case reports or series published between 1930 and 2019. In total, 250 cases of plague were described from seven countries: China (n=114) [26–28], the United States (n=96) [8,29–35], Libya (n=17) [36], Kazakhstan (n=12) [37], Madagascar (n=9) [22], South Africa (n=1) [38], and Saudi Arabia (n=1) [39]. More males were infected, with a wide age range from 1 to 69 years. There were 125 cases diagnosed with primary bubonic plague (mostly axillary buboes), 70 with primary pneumonic plague, eight with primary septicaemic plague and two with primary intestinal plague.

#### Risk of bias

Most of the included studies adequately described the main characteristics of participants (Table 3, details in S7 Appendix). Efforts to trace all contacts from the index case was unknown for 12 studies. This means that there was no information on whether other persons were exposed to the source of infection without getting infected, which makes judgment on assessing contagiousness of cadavers difficult. In eight studies, laboratory methods used for defining confirmed cases of plague were unknown or partial for most of the cases described by each study. In these cases, plague diagnosis was however highly suspected due to clinical and epidemiological data. In 11 studies, the route of transmission and the cause-effect from the source of infection to the infected person were highly plausible. In the remaining studies, while the source and route of transmission can eventually be plausible, there was a lack of information to make this judgment, and in some case series, transmission via fleas might not have been fully excluded for all cases.

**Table 3.** Risk of bias summary for studies on plague acquired from human or animal cadavers (part 2)

| Study ID | Were patient characteristics adequately reported? | Was there some effort to trace all contacts from the index case? | Were the methods used for tracing contacts adequate? | Were the laboratory methods used for defining a confirmed case of plague reliable? | Was the route of transmission plausible? | Was the cause-effect of transmission plausible? |
| --- | --- | --- | --- | --- | --- | --- |
| CDC 1992 | Yes | Unknown | Not applicable | Yes | Yes | Yes |
| Christie 1980 - case series 1 | Partial | Unknown | Not applicable | Partial | Yes | Yes |
| Christie 1980 - case series 2 | Partial | Unknown | Not applicable | Partial | Partial | Partial |
| **Gage 2000** | Yes | Unknown | Not applicable | Yes | Yes | Yes |
| **Ge 2015 - case report** | Yes | Yes | Yes | Yes | Yes | Yes |
| **Ge 2015 - case series** | Partial | Unknown | Not applicable | Unknown | Partial | Partial |
| **Kartman 1960** | Partial | Unknown | Not applicable | No | Yes | Yes |
| **Kartman 1970** | Partial | Unknown | Not applicable | Unknown | Yes | Yes |
| **Kugeler 2015** | No | Unknown | Not applicable | Unknown | Partial | Partial |
| **Mitchell 1930** | Yes | Unknown | Not applicable | Unknown | Yes | Yes |
| **Poland 1973** | Yes | Yes | Yes | Yes | Yes | Yes |
| **Ratsitorahina 2000** | Yes | Yes | Unknown | Yes | Yes | Partial |
| **Saeed 2005** | Yes | Yes | Yes | Yes | Yes | Yes |
| **Sagiev 2019** | No | Unknown | Not applicable | Unknown | Unknown | Unknown |
| **Von Reyn 1976** | Yes | Yes | Yes | Yes | Yes | Yes |
| **Wong 2009** | Yes | Yes | Yes | Yes | Yes | Yes |
| **Wu 2009** | Yes | Unknown | Not applicable | Yes | Yes | Partial |
| **Zhang 2007** | Partial | Unknown | Not applicable | Partial | Unknown | Partial |

#### Findings

Human cadavers were the source of exposure in two studies including 10 cases [22,38] and in a third study of 32 cases with no disaggregated data between contact with alive humans and human cadavers [28]. In the remaining studies, the exposure was animal cadavers including camels, goats, cats, bobcat, fox, coyote, mountain lion, Tibetan sheep, marmots, dogs, rabbits, squirrels, and other rodents. In most cases, the type of exposure consisted of activities related with the animal cadaver such as killing the animal, skinning the cadaver, or performing autopsy of the infected animal, which require a relatively long and close exposure with the source of infection. It was not possible to disaggregate data on the cause of transmission between handling or consuming the infected animal in three studies (56 cases) [27,28,36], to affirm whether plague transmission was due to the contact with a human or animal cadaver or to any alternative routes in four studies (45 cases) [22,28,29,36] and to ascertain whether plague was transmitted from a cadaver or from interhuman contact in three studies (44 cases) [22,28,36].

Only one study directly specified the duration of time between the death of the infected animal and the start of exposure, being around 35 hours [8]. Three studies gave enough details to affirm that the time of exposure was within 24 hours following the death of the infected animal [22,34,39]. In 26 additional cases from three studies, we can deduce an immediate exposure from infected animals that were just killed by the cases who developed plague [31,32,36]. Among cases with bubonic plague, five cases from four studies had open skin lesions while handling the cadaver with bare hands [32,34,35,39]. Other persons who had no skin lesions were exposed to the same source of infection but were not infected [34,35]. While another study associated the route of transmission to “direct contact from infectious body fluids of the cat cadaver” [30], there is no further details of the entry point of the plague bacilli in the remaining cases, other than “direct handling” of the cadavers. Most cases of bubonic plague were axillary, which is consistent with the inoculation of *Y. pestis* through cuts in the hands or arms. Among cases who developed primary pneumonic plague, one study attributed transmission via inhalation of aerosols generated while handling the cadaver during the necropsy, and another study mentioned “exposure to aerosols” [8,26]. A third study attributed the pneumonic infection as a result of “active participation in the funeral ceremonies” without providing further details [22].

#### Summary

Direct skin contact with blood could cause bubonic and septicaemic plague. The risk is increased in people with cuts or skin abrasions. We do not know about infectiousness of other body fluids. Potentially pneumonic plague could be transmitted by actions that provoke aerosolization of infected body fluids, but these will require considerable manipulation of the cadaver.

### 3) Infectiousness of body fluids of animal or human cadavers, including length of cadaver infectiousness

We identified two studies that detailed the length of infectiousness of plague-infected animals and none in human cadavers. One experimental study conducted in Madagascar five decades ago aimed to isolate *Y. pestis* from rodent cadavers that died from septicaemic plague and were buried in laterite alone or in laterite enriched with manure to simulate local conditions [40]. *Y. pestis* was successfully isolated after five and ten days but failed to be isolated at 15 days after death of the rodents. The second study, already described above for objective 2, reported the case of a wildlife biologist who has been in contact with a mountain lion around 35 hours after the death of the animal [8]. The time of death was identified from a ‘mortality signal’ transmitted from the animal’s radio-collar after six hours of no movement. *Y. pestis* was isolated by culture and subtyped by pulsed-field gel electrophoresis from the animal’s tissues. The same strain was isolated from the biologist, supporting the mountain lion as the source of the biologist’s infection. Both studies were judged to be at low risk of introducing bias.

In summary, we do not know for how long *Y. pestis* can survive in body fluids of people that die from plague, and we thus do not know for how long the cadaver is contagious. One case of infection from an animal 35 hours after death means whatever the size of the risk, it may well extend beyond 24 hours.

## Discussion

Pandemics of plague reported historically suggest that the risk of human-to-human transmission in this context is high for pneumonic plague but this has been contested.

Indeed, a systematic review summarized data from historical records and contemporary experience, and based on qualitative analysis concluded that “pneumonic plague is not easily transmitted from one person to another” [41]. Quantitative assessment of transmissibility of plague has also been performed, using mathematical models based on historical data [42,43]. Our data, which include most recent outbreaks (older reports usually do not provide enough information to allow inclusion in the review) support that pneumonic plague is transmissible from human to human, sometimes only after close and prolonged exposure. Historical records that were not eligible for this review constitute a useful additional source of information related to transmissibility of pneumonic plague. Some of them reported experiments that demonstrated the isolation of *Y. pestis* from sputum of people sick with pneumonic plague [44,45].

Looking at the evidence gathered in this review, respiratory secretions consisting of blood-stained sputum were clearly reported as the source of plague transmission. From studies describing plague acquired from cadavers, the type of contaminated body fluids causing plague transmission was not clearly described, although it could mostly be assumed to be blood. Indeed, the activities described to be the cause of infection included skinning of the animal, cutting carcasses, flaying, post-mortem examinations, all of them involving contact with blood. However, it could potentially be linked with other body fluids infected such as urine, faeces, gastric content or bubo pus. We did not find evidence that the disease can be transmitted by body fluids other than sputum and blood. In addition, we do not know for how long *Y. pestis* survive in the body fluids, and for how long the cadaver is contagious. From real life situations, we found only one study describing plague transmission from an animal that was dead for around 35 hours before the first contact with the infected human.

The included studies described two main routes of transmission. The first one is the inhalation of particles, which can result in pneumonic plague. From alive sick persons, contaminated droplets are generated by cough associated with bloody sputum. From cadavers, contaminated droplets will no longer be produced by cough but can be generated from body fluids – mainly blood – by handling the cadaver, for example when performing a necropsy or when preparing body for funerals. In any case, a close and prolonged exposure is probably needed for transmission of the disease. The second route of transmission is through handling cadavers, described for prolonged exposure involving invasive procedures. Skin cuts or abrasion in the hands are described in some of the persons who got infected. In other studies, this was not commented on. Thus, it is difficult to know whether transmission through intact skin can occur, although from first principles this seems unlikely. We did not find any study describing plague acquired through contact with mucosa.

In some cases, it is difficult to discriminate between the different routes of transmission from plague-infected human cadavers (body fluids, clothing contaminated with body fluids, or fleas). In part 1, we showed that human-to-human transmission was possible. All infected cases presented primary pneumonic plague, which suggests inhalation of particles to be the source of infection, while fleas were very unlikely to be involved. In part 2, we reported plague transmission from human and animal cadavers. Most cases were described from animals, with no involvement of potential contaminated clothing in the transmission of the disease. It is difficult to totally exclude the possibility that some of the infected cases presenting with bubonic plague were contaminated by fleas. While most cases were contaminated from animals with no involvement of clothes that could contain fleas, there could have been fleas on the corpses. However, our inclusion criteria limited the likelihood of plague transmission by fleas and we appraised the plausibility of the route of transmission (human to human or from corpses) for each included study. We excluded cases related with flea or unknown transmission [29] and highlighted when there was evidence of absence of flea bite [32], or at contrary, when transmission via flea might not have been fully excluded [33].

In conclusion, there is risk of plague transmission from human cadavers (Fig 4). Inhalation of respiratory droplets produced by intense manipulation of the cadaver could result in pneumonic plague, probably following close and prolonged exposure. Direct skin contact of infected body fluids (blood but unclear for other body fluids) could cause bubonic plague and eventually septicaemic plague when the persons present cuts in their hands.

**Fig 4.**
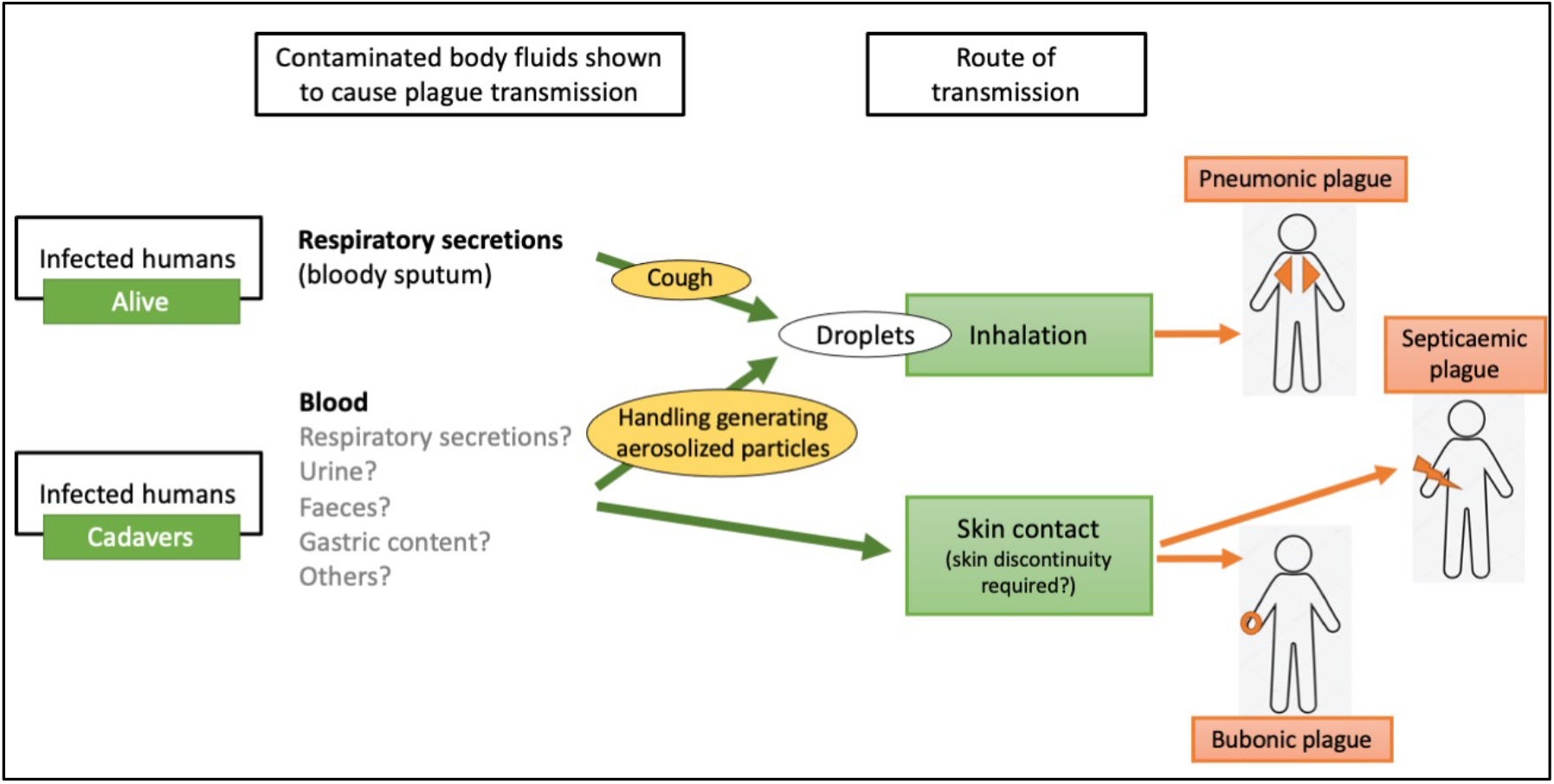
Summary of the transmission routes evidenced by the literature gathered in this review.

## Data Availability

This is a systematic review of published studies that can be found in the public domain.

## Acknowledgments

We acknowledge Xin Wang for retrieving the full text manuscripts in Chinese, for summarizing the Chinese manuscripts so that SJ and NS could assess eligibility of the study, and for conducting data extraction of the two included Chinese studies. We also acknowledge Vladimir Dubyanskiy for retrieving some of the full text manuscripts in Russian and for providing a brief summary of the Russian papers in order to assess eligibility. We are grateful to Vittoria Lutje, the Information Retrieval Specialist of the Cochrane Infectious Diseases Group for her help with the literature search strategy and for conducting the search.

## Supporting information

**S1 Appendix. Search strategy**

**S2 Appendix. Characteristics of excluded studies S3 Appendix. Adapted quality appraisal tool**

**S4 Appendix. Summary of studies describing infectiousness of different body fluids of people ill with plague (review part 1)**

**S5 Appendix. Characteristics of included studies for infectiousness of different body fluids of people ill with plague (review part 1)**

**S6 Appendix. Summary of studies describing plague acquired from human and animal cadavers (review part 2)**

**S7 Appendix. Characteristics of included studies for plague acquired from human and animal cadavers (review part 2)**

