## Appendices for "Risk of plague transmission from human cadavers: a systematic review"

**S1 Appendix. Search strategy**

1     [plague.mp](http://plague.mp/). or *plague/

2     yersinia [pestis.mp](http://pestis.mp/). or Yersinia pestis/

3     1 or 2

4     (transmission or transmitted).ab. or (transmission or transmitted).ti.

5     disease [transmission.mp](http://transmission.mp/). or disease transmission/

6     (contamination or contaminated or spread*).ab. or (contamination or contaminated or spread*).ti.

7     (infection or infectious).ab. or (infection or infectious).ti.

8     4 or 5 or 6 or 7

9     3 and 8

10     cadaver/ or cadaver*.mp.

11     corpse*.mp. or corpse dismemberment/

12     remains.ab. or remains.ti.

13     carcass*.ab. or carcass*.ti.

14     body fluid*.ab. or body fluid*.ti.

15     posthumous care/ or burial/ or burial*.mp.

16     (cemetery or cemeteries).mp. [mp=title, abstract, heading word, drug trade name, original title, device manufacturer, drug manufacturer, device trade name, keyword, floating subheading word, candidate term word]

17     funeral*.mp.

18     [entombment.mp](http://entombment.mp/).

19     10 or 11 or 12 or 13 or 14 or 15 or 16 or 17 or 18

20     9 and 19

**S2 Appendix. Characteristics of excluded studies**

| **Study ID** | **Reason for exclusion** |
| --- | --- |
| Aikimbajev 2003 | Transmission of plague via fleas and slaughter with no further information from the cadavers. |
| Ainiwaer 2011 | Report of the case of *Y. pestis* strain isolated from a cadaver of rodent, with no details on the time the rodent died before the isolation of *Y pestis*, and with no subsequent infection of human plague. |
| Arbaji 2005 | Cases of plague by consumption of raw meat from an infected camel. |
| Asaku 2016 | Abstract. No report of new case of plague. Rodent carcasses with no specification of isolation of *Y pestis* by culture nor time of plague positivity from death. |
| Baltazard Bahmanyar 1960 | Description of interhuman transmission via parasites. |
| Baltazard Seydian 1960 | Description of interhuman transmission via parasites. |
| Bannerman 1906 | This manuscript describes the spread of plague: a) from man to man directly; b) by means of animals; c) by means of clothes; but also by grain, by air, by water, among others. Although this historical piece of work adds valuable information, not enough details are provided for inclusion in the review. |
| Biggins 2019 | Study on maintenance of disease among rodents. |
| Boegler 2018 | This is a rodent carcass surveillance programme. Carcasses were tested by direct fluorescent antibody against F1 Ag, with no description of direct plague transmission to humans nor identification of *Y. pestis* from the carcasses by culture, and duration from death is not described. |
| Boisier 2002 | Epidemics of bubonic plague in Madagascar with no direct information on method of interhuman transmission. |
| Boone 2009 | Study of carcass removal by carnivores with finding of uninfected carcasses. |
| Chalmers 1900 | No clear data on human-to-human transmission of plague. |
| Cohn 2018 | Chapter of an history book. Historical aspects of changes in burial rituals with epidemics discussed, but no direct evidence on transmission via cadavers. |
| Danforth 2016 | Two new cases of plague, for which transmission was found to be related with rodents and fleas (no direct transmission). |
| Davis 2007 | Plague transmission within burrow systems. |
| Didelot 2017 | Description of plague transmission from cadavers using mathematical modelling with no description of real cases with direct transmission. |
| Easterday 2012 | Study of genetic changes in plague bacilli. |
| Esposito 1992 | Although the paper mentions “plague has been reported in individuals who have come into contact with the infected carcasses of deer, antelope, foxes, bobcats and coyotes” no further details are provided that would allow inclusion of this manuscript. Reference cited for this statement was checked. |
| Evans (B) 2018 | Letter to the editor on transmission via respiratory droplets of parasites. |
| Ganière 2001 | Study looking at zoonoses transmitted by cats and dogs. There were no cases of plague described transmitted from dead cats or dogs, but only by bites and scratches. |
| George 1941 | Description of transmission to rodents via parasites with no details on persistence or infection from rodent cadavers. |
| Gimlette 1909 | Description of plague outbreak describing 31 persons infected, including pneumonic and bubonic plague. Mode of transmission is unclear; infection might have been brought from imported clothing with fleas, and report of rats migration and fleas. |
| Graf 2006 | This paper looked at simulation of scenario in case of death of person with highly contagious disease. |
| Healing 1995 | Description of infection hazards of human cadavers from both old interments and the recently deceased. There were no cases of plague transmission described from the recently deceased. |
| Jellison 1939 | Study of bird infection when eating infected rodents. However, experiments on whether viable organisms were present in the faeces of birds being fed with plague-infected guinea pig tissue (not natural infection). |
| Krishnaswami 1972 | Investigation of the presence of wild plague foci in an area where an outbreak took place, carrying out serological studies from the reported outbreaks and determining the prevalent rodent and flea fauna in the area. No direct evidence on mode of transmission for plague cases was reported. |
| Kuznetsov 2018 | Cartography applied for natural foci of plague. |
| Lynteris 2018 | Chapter of an history book. Historical aspects of changes in burial rituals with epidemics discussed, but no direct evidence on transmission via cadavers. |
| Madras 1917 | Report on veterinary services with no description on plague transmission. |
| Matsuo 1912 | Case of human infected from his donkey that died five days before the onset of disease. No clear mode of transmission described and contact with the donkey was established before the donkey died. |
| Mayevsky 1999 | Description of transmission of plague among sousliks by fleas through winter. |
| Mitra 1907 | Description of a plague outbreak with no laboratory confirmation of the disease. The author raises the possibility of transmission from a dead body; however, this remains uncertain, as reported by the author: ‘How this man contracted plague is a mystery’; ‘I have heard a story’; ‘There are two probable stories’. |
| Nishiura 2006 | Mode of transmission not described. |
| Njunwa 1989 | Study on rodents and parasites with no description of timing for detection of bacilli from rodents. |
| Nyirenda 2017 | This study aimed to investigate potential risk factors that are possibly associated with facilitating, maintaining, and transmitting sylvatic and murine plague to humans in the study area. |
| Poleykett 2018 | Review on rituals on burial of dead bodies with no evidence on transmission via cadavers. |
| Reed 1970 | There are cases of bubonic plague reported from dead rodents to humans but attributed to transmission via fleas. |
| Reynolds 2011 | Description of humans killing of animals to prevent spread of zoonoses, with no evidence provided on transmission of plague from animals. |
| Richgels 2016 | Simulating plague transmission through animal carcasses, with no description of direct cases. |
| Rollo-Koster 2018 | Chapter of an history book. Historical aspects of changes in burial rituals with epidemics discussed, but no direct evidence on transmission via cadavers. |
| Russo 1930 | Study based on experimental investigation of plague in insects. |
| Simpson 1909 | Full text manuscript could not be retrieved. |
| Sludsky 2018 | Description of the large-scale epidemics of plague around the world, but no description of any case of human plague attributed to animal or human cadavers. Mention of cases of pneumonic plague probably transmitted by humans, with no further details. |
| Sotnikov 1973 | Full text manuscript could not be retrieved. |
| Stepanov 1990 | Description of human plague in 1988 in the Republic of Kazakhstan, considering the human infection by fleas. |
| Strobel 2004 | Laboratory techniques with no description of cases of plague transmission. |
| Strong 1912 | Experiments around plague transmission with no description of human-to-human plague transmission. |
| Suchkdv 1965 | Full text manuscript could not be retrieved (It pertains to a book located in a single library in Russia that does not participate in interlending). |
| Teh 1923 | Experiments and additional details on the role of rodents in plague. No description of human-to-human plague transmission. |
| Titus 2016 | Abstract for conference on active surveillance with no information provided on mode of transmission. |
| Van Arsdel 1987 | Letter to the editor. |
| Vasin 2014 | Historical narrative of cases of plague with no direct description of cases. |
| Walsh 2015 | Modelling on transmission of plague among animals, with no direct cases of plague transmission to animals via cadavers reported. |
| Webb 2006 | No cases describing transmission via animal cadavers. |
| Werner 1984 | Transmission through direct contact with alive cat, which ended dying. |
| Whittles 2016 | Transmission of plague using models on historical data. Cases are not confirmed. In this model human-tohuman transmission incorporates direct transmission and transmission via parasites together. |
| Zhongliang 2016 | Historical narrative manuscript on a doctor involved in plague. |
| Zou 2005 | Description of a bubonic outbreak in China, with no evidence of inter-human transmission. |
| Several authors 1912 | Report of the international plague conference held at Mukden in 1911. This report is a ‘well-printed book of about 500 pages’, to which we could not access (first page available only). |

**S3 Appendix. Adapted quality appraisal tool**

| *For the following questions, tick answer in one of the columns.* | Yes | Partial | No | Not applicable | Notes |
| --- | --- | --- | --- | --- | --- |
| 1. Were patient characteristics adequately reported? |  |  |  |  |  |
| 2. Was there some effort to trace all contacts from the index case? |  |  |  |  |  |
| 3. Were the methods used for tracing contacts adequate? |  |  |  |  |  |
| 4. Were the laboratory methods used for defining a confirmed case of plague reliable? |  |  |  |  |  |
| 5. Was the route of transmission described plausible? |  |  |  |  |  |
| 6. Was the cause-effect of transmission plausible? |  |  |  |  |  |

**S4 Appendix. Summary of studies describing infectiousness of different body fluids of people ill with plague (review part 1)**

| **Study ID** | **Setting**  **Date** | **Index case** | **Infected contacts** | **Non-infected contacts** | **Contagiousness** | **Contaminated body fluids and route of transmission (as per study authors)** |
| --- | --- | --- | --- | --- | --- | --- |
| Almeida 1981 (16) | Brazil  1977-1979 | 1 (BP) | 1 (PPP) | Not reported | Not reported | Not reported |
| Begier 2006 (24) | Uganda  2004 | 2 (SPP) | 2 (PPP) | 23  (no PEP) | Attack rate 8% | ‘Respiratory droplets rather than aerosols’ |
| Bertherat 2011 (25) | DRC  2004-2005;  2006 | Not reported | 292 (290 PPP, 2 SP) | Not reported | Not reported | Not directly stated, assumed from sputum and ‘aerosolized bacteria spread through coughing’ |
| Evans 2018 (17) | South Africa  1904 | Unable to be traced | 121 (113 PP, 2 SP, 6 mixed) | Not reported | R_t_: peak of 2-4 | ‘There is little evidence to confirm the conventional view that such cases originated through airborne transmission from patients with bubonic plague in whom secondary pneumonic plague had developed (mixed cases).’ |
| Kellogg 1920 (19) | US  1919 | 1 (BP; SPP) | 13 (PPP) | Not reported | Not reported | ‘Droplet infection and personal contact’ |
| Kugeler 2015 (18) | US  1900-1925 | Not reported | 49 (PPP) | Not reported | Not reported | ‘Human-to-human transmission’ |
| Rabaan 2019 (20) | Madagascar  2017 | 1 (SPP) | 1861 (PPP)* | Not reported | Not reported | ‘Via respiratory droplets’ |
| Ramasindra-zana 2017 (21) | Madagascar  2015 | 1 (SPP) | 13 (PPP) | 123 from index case  (PEP in 35) | R_0_: 1.44; TR: 0.41 | ‘The matched genetic grouping between the 2 human samples is consistent with human-to-human transmission.’ |
| Ratsitorahina 2000 (22) | Madagascar  1997 | 1 (SPP) | 17 (PPP) | 154  (PEP) | Attack rate 8.4% | ‘Direct transmission of *Y. pestis* through infective cough droplets’ |
| Richard 2015 (23) | Madagascar 2011 | 1 (PP) | 19 (PPP) | 41  (PEP in 39) | Attack rate 55% | Not directly stated, assumed ‘pathogen is transmitted as an aerosol by droplets or by contaminated dust’ |

Abbreviations: BP: bubonic plague; DRC: Democratic Republic of the Congo; PEP: post-exposure prophylaxis; PP: pneumonic plague; PPP: primary pneumonic plague; R_0_: reproductive number; R_t_: estimation of time varying, which is the average number of secondary infections resulting from an infectious person; SP: septicaemic plague; SPP: secondary pneumonic plague; TR: transmission rate, in susceptibles per day.

^*^2417 cases of plague were registered during this outbreak, including both pneumonic and bubonic forms of plague. In another manuscript describing the same outbreak, authors mentioned that pneumonic plague was confirmed in 77% of them (10). We therefore report here the number of patients reported with pneumonic plague.

**S5 Appendix. Characteristics of included studies for infectiousness of different body fluids of people ill with plague (review part 1)**

| **Almeida 1981** | |
| --- | --- |
| **Basic information** | **Setting:** Brazil, states of Ceara, Pernambuco, and Paraiba  **Date:** December 1977 to May 1979  This manuscript reported the cases of plague in Brazil during a two-year period. The other cases described are bubonic cases, and no other case can be ascertained to be associated to human to human transmission. |
| **Index cases** | **Number of index cases:** 1  **Form of plague:** bubonic plague, with no further details.  **Description of infected cases:** Whether the case developed respiratory symptoms is not reported. It is however mentioned that none of the cases with confirmed plague got severely ill. |
| **Infected cases from index cases** | **Number of contacts infected:** 1  **Form of plague:** primary pneumonic plague  **Description of infected contacts:** 33-year-old female, family contact of the index case. Absence of bubo, but signs of pneumonia.  **Route of transmission** (as described by study authors): not reported |
| **Plague diagnosis** | **Definitions of plague cases:** not reported  **Laboratory findings:** the infected case presented a positive sputum culture for *Y pestis* and positive serology during convalescence. |
| **Contacts** | **Number of contacts:** not reported  **Attack rate**: not reported  **Method of contact tracing:** not reported  **Definition of ‘contact’:** not reported  **Other relevant data describing contacts:** none |
| **Other relevant notes** | None |

| **Quality appraisal** | **Authors’ judgment** | **Support for judgment** |
| --- | --- | --- |
| 1. Were patient characteristics adequately reported? | Partial | Some data are given for the infected contact, but no information is provided on the case index other than bubonic plague. |
| 2. Was there some effort to trace all contacts from the index case? | Unknown | Not reported |
| 3. Were the methods used for tracing contacts adequate? | Not applicable | Not applicable |
| 4. Were the laboratory methods used for defining a confirmed case of plague reliable? | Yes | The diagnosis of the infected contact was confirmed by a positive sputum culture for *Y pestis* and positive serology during convalescence. |
| 5. Was the route of transmission plausible? | No | The index case is a case of bubonic plague with no other details provided. The infected case is a primary pneumonic plague. Although we can assume that the index case developed secondary pneumonic plague, this is not mentioned. Also, authors said ‘none of the cases with confirmed plague got severely ill’ |
| 6. Was the cause-effect of transmission plausible? | Unknown | Following the above statement, unknown what was the route of transmission considered to judge the plausibility of the cause-effect transmission. |

| **Begier 2006** | |
| --- | --- |
| **Basic information** | **Setting:** Uganda  **Date:** December 2004 |
| **Index cases** | **Number of index cases:** 2 (case A and case B)  **Form of plague:** secondary pneumonic plague (primary bubonic plague in A, unknown lymphadenopathy in B but according to evolution of the disease)  **Description of infected cases:**  No social link found between both cases and no contact in the week prior the onset of the disease.  Developed productive cough progressing to grossly bloody sputum  Survived more than 1 week without appropriate treatment, therefore severely ill towards the end, before fatal outcome. |
| **Infected cases from index cases** | **Number of contacts infected:** 2  **Form of plague:** primary pneumonic plague  **Description of infected contacts:** the primary caregiver and mother of case (A), and the primary caregiver and sister of case (B)  **Route of transmission** (as described by study authors): ‘respiratory droplets, rather than aerosols’ |
| **Plague diagnosis** | **Definitions of plague cases: ‘**probable pneumonic plague case’: respiratory illness of acute onset with cough producing grossly bloody sputum. ‘Definite pneumonic plague case’: probable case with laboratory evidence of plague infection.  **Laboratory findings:** 3 cases already buried when plague outbreak identified. One case with PCR positive in sputum, culture negative, positive immunochromatography |
| **Contacts** | **Number of contacts:** 25  **Attack rate**: 8%  **Method of contact tracing:** Prospective active surveillance in the affected region. Additional retrospective pneumonic plague surveillance by interviewing private drug shop owners, business owners, traditional healers, and other area residents.  **Definition of ‘contact’:** ‘Close contact’ with any of the index case (i.e., touched) after onset of cough productive of bloody sputum and before death.  **Other relevant data describing contacts:**  The contacts did not receive antimicrobial prophylaxis as ‘more than a week had passed since the index patients’ deaths when the outbreak was reported.’  Non-infected contacts included 3 family members who slept in the same bed as infected cases the night before the death of the infected cases.  Non-infected contacts also included several persons who slept with their heads at a distance <2 m from the coughing plague patient.  ‘In addition, around 200 people attended the 2 cases’ funeral and around 75 persons touched the blanket that wrapped one index patient’s body, the same blanket that was used during the patient’s final days of illness. No contacts used masks, gloves, or any other form of respiratory protection.’  ‘No additional pneumonic plague cases were identified during December and in the weeks after the outbreak report. However, through active surveillance we identified 3 probable bubonic plague patients who came to the subcounty’s local health center in the first half of January, an increase from a baseline of 0 cases per month in the preceding 3 months.’ |
| **Other relevant notes** | ‘Our patients’ clinical course provides clues to why pneumonic plague patients usually infect few persons and why, for example, an air travel–associated outbreak would be unlikely. Our case-patients were visibly short of breath, coughing grossly bloody sputum, and barely ambulatory before transmitting the disease. Thus, when patients are substantially contagious, they are unlikely to be traveling by air and, if so, would appear ill enough to alarm nearby passengers. In most settings, persons this ill are at home or in the hospital. Recent reviews support this observation because most reported pneumonic plague transmissions involve family, friends, or medical professionals caring for ill persons at home or in the hospital.’ |

| **Quality appraisal** | **Authors’ judgment** | **Support for judgment** |
| --- | --- | --- |
| 1. Were patient characteristics adequately reported? | Yes | Form of plague and relationship between index cases and contacts are clearly reported |
| 2. Was there some effort to trace all contacts from the index case? | Yes | Contacts are well defined, traced and described. |
| 3. Were the methods used for tracing contacts adequate? | Yes | Prospective active surveillance and retrospective pneumonic plague surveillance conducted through interviews. |
| 4. Were the laboratory methods used for defining a confirmed case of plague reliable? | Partial | 3 cases were already buried and therefore no samples were analyzed.  1 case was confirmed by PCR and immunochromatography |
| 5. Was the route of transmission plausible? | Yes | Inhalation of infected ‘respiratory droplets’ |
| 6. Was the cause-effect of transmission plausible? | Yes | Both index cases are described to develop cough and ‘grossly bloody sputum’ |

| **Bertherat 2011** | |
| --- | --- |
| **Basic information** | **Setting:** Democratic Republic of the Congo. The 2005 outbreak occurred in a diamond mining camp in a remote area with no previous cases of plague reported in the area. The 2006 outbreak occurred in a gold mining camp.  **Date:** first outbreak from December 2004 until March 2005; second outbreak from August to November 2006 |
| **Index cases** | **Number of index cases:** not reported  **Form of plague:** not reported  **Description of infected cases:** not reported |
| **Infected cases from index cases** | **Number of contacts infected:** 130 (2005 outbreak) and 162 (2006 outbreak)  **Form of plague:** 128 pneumonic cases + 2 septicemic cases from the 2005 outbreak  **Description of infected contacts:** not reported  **Route of transmission** (as described by study authors): not reported |
| **Plague diagnosis** | **Definitions of plague cases:** WHO case definitions  **Laboratory findings:** suspicion based on the clinical evolution and the outbreak characteristics. Once WHO team was on site, microbiology testing from blood and sputum samples, immunochromatography form sputum samples, and serology from paired serum samples. Based on the WHO case definitions: 5 confirmed, 10 probable and 115 suspected cases from the 2005 outbreak, and 23 confirmed, 22 probable and 117 suspected cases from the 2006 outbreak. |
| **Contacts** | **Number of contacts:** not reported  **Attack rate**: not reported  **Method of contact tracing:** through WHO intervention  **Definition of ‘contact’:** not reported  **Other relevant data describing contacts:** close contacts received chemoprophylaxis |
| **Other relevant notes** | ‘Pneumonic plague is of serious concern because of the potential for human- to-human transmission from aerosolized bacteria spread through coughing. Pneumonic plague can lead to localized outbreaks, or even devastating epidemics, because the infectious dose by inhalation can be as low as 100–500 organisms’ |

| **Quality appraisal** | **Authors’ judgment** | **Support for judgment** |
| --- | --- | --- |
| 1. Were patient characteristics adequately reported? | No | Description of index cases and infected contacts not reported other than number of forms of plague of infected contacts. |
| 2. Was there some effort to trace all contacts from the index case? | Partial | Although the index case is not reported, contacts were traced from the moment the WHO team reached the outbreak setting and put in place adequate measures. |
| 3. Were the methods used for tracing contacts adequate? | Unknown | Not reported. |
| 4. Were the laboratory methods used for defining a confirmed case of plague reliable? | Yes | Cases were defined as confirmed, probable and suspected cases of plague according to the WHO definitions. |
| 5. Was the route of transmission plausible? | Yes | Although the index case is not reported, transmission among the rest of the cases causing the outbreak is assumed to be ‘human to human transmission from aerosolized bacteria spread through coughing’. |
| 6. Was the cause-effect of transmission plausible? | Yes | Context of pneumonic plague outbreak following the route of transmission described above. |

| **Evans 2018** | |
| --- | --- |
| **Basic information** | **Setting:** South Africa, Johannesburg  **Date:** January to March 1904  The study authors based their manuscript on ‘the Rand Plague Committee’ (the RPCR), a report ‘that documented the principal findings together with the data on which their inferences were based’. |
| **Index cases** | **Number of index cases:** unknown  **Form of plague:** unknown  **Description of infected cases:** “The investigators were unable to trace the index case(s) and could only speculate that ‘Plague infected rice was imported from Bombay during December, 1903 and January 1904. From this rice a few Indians were infected with the pneumonic form of plague.’” |
| **Infected cases from index cases** | **Number of contacts infected:** 121  **Form of plague:** 113 primary pneumonic plague, 6 mixed, 2 septicemic plague. The manuscript also described 40 cases of bubonic plague that were attributed to a ‘low-key epizootic of rats’ (not included for the purpose of this review)  **Description of infected contacts:** Mainly Indians, but also ‘whites’, ‘natives’ and ‘coloureds’. Both sex affected, high proportion of males. Survival for 31 bubonic plague and 2 pneumonic plague.  Before death, patients with primary pneumonic plague, presented with some ‘scanty but blood-stained expectorations.’  **Route of transmission** (as described by study authors):  ‘Investigations of the Indian community identified 16 probable transmissions involving nursing, preparing bodies for funerals, attending funerals, or close family members.’ ‘transmission seemed to follow relationship pathways involving intimate contact.’  ‘There is little evidence to confirm the conventional view that such cases originated through airborne transmission from patients with bubonic plague in whom secondary pneumonic plague had developed (mixed cases) and no evidence that a person from outside Johannesburg introduced pneumonic plague into the area.’ |
| **Plague diagnosis** | **Definitions of plague cases:** ‘Pure pneumonic cases were those in which no buboes could be found, but in which there was definite bronchopneumonia. The mixed cases were those in which there was definite bronchopneumonia, as well as buboes, and the *B. Pestis* [sic] was recovered both from the foci in the lungs and from the bubo. The septicaemic cases were those without either signs of pneumonia or buboes.’  **Laboratory findings:** ‘*Y. pestis* in samples of sputum or tissues from organs including the lung, spleen, and liver. Bacteria were cultured, identified morphologically, and subsequently confirmed by inoculation into rabbits and guinea pigs.’ |
| **Contacts** | **Number of contacts:** unknown  **Attack rate**: not calculated. Estimation of time varying (R_t_), which is the ‘average number of secondary infections resulting from an infectious person’, reaching value of 2-4.  **Method of contact tracing:** ‘inspectors were appointed to search for additional sick persons’  **Definition of ‘contact’:** not reported  **Other relevant data describing contacts:**  ‘The decrease in estimated transmissibility coincides with the start of the isolation process on March 18, suggesting that this strategy was probably effective.’  ‘A particularly noteworthy aspect of this outbreak of primary pneumonic plague was that none of the 9 escapees from the Coolie Location transmitted the disease to the wider population; the RPCR also lists lack of transmission by many other case-patients.’’ Nevertheless, within social networks characterized by family connections, employment, caste, and so on, the disease spread rapidly.’ |
| **Other relevant notes** | ‘Transmission rates rapidly diminished after implementation of control measures, including isolation and safer burial practices’  ‘As the outbreak progressed, most deaths occurred in hospitals that allowed some control of burial practices. The RCPR states that “... in the case of Hindoos and the Mohammedans [sic]. The former were allowed to bury their dead: the latter, who have certain religious functions to perform were given a room in the mortuary to perform the rite. They were warned of the dangers of handling the cadavers, and it was suggested to them that the washing should be performed with a solution of corrosive sublimate.”’  ‘pattern in which the disease is transmitted to relatives, friends, or caregivers but not to more loosely associated contacts’  ‘It is well known that primary pneumonic plague rapidly incapacitates the patient, who is then incapable of reaching potential contacts within the most infectious period. Nevertheless, this study shows that relatively high rates of transmission were achieved in Johannesburg in 1904, as demonstrated by the peak values for the estimated time-varying Rt.’ |

| **Quality appraisal** | **Authors’ judgment** | **Support for judgment** |
| --- | --- | --- |
| 1. Were patient characteristics adequately reported? | Yes | Although the index case could not be traced, characteristics of the infected contacts are well reported. |
| 2. Was there some effort to trace all contacts from the index case? | Unknown | Not reported. |
| 3. Were the methods used for tracing contacts adequate? | Not applicable | Not applicable |
| 4. Were the laboratory methods used for defining a confirmed case of plague reliable? | Yes | Isolation of *Y pestis* from biological samples |
| 5. Was the route of transmission plausible? | Yes | Although the index case was not traced, transmission among the rest of the cases causing the outbreak is assumed to be transmission between humans with pneumonic plague. |
| 6. Was the cause-effect of transmission plausible? | Yes | Context of pneumonic plague outbreak following the route of transmission described above.  Cases of bubonic plague were also reported, and attributed to flea and rats associated transmission. |

| **Kellogg 1920** | |
| --- | --- |
| **Basic information** | **Setting:** United States, Oakland  **Date:** August to September 1919 |
| **Index cases** | **Number of index cases:** 1 (case A)  **Form of plague:** bubonic plague with secondary pneumonic plague.  **Description of infected cases: m**ale who went hunting 2 and 4 days before developing symptoms: fever, chest pain and right axillary bubo. Death at day 6 from start of symptoms. |
| **Infected cases from index cases** | **Number of contacts infected:** 13 in total. One case (B) from the index case (A), 12 additional cases from contacts with case B (5 (C-G) from case B and further 7 (H-N) from three of the subsequent contacts).  **Form of plague:** primary pneumonic plague  **Description of infected contacts:** adults, 9 males, 4 females. Direct close contact described for each of them, such as visit, nurse, physician, or living with an infected person. Only one survived.  **Route of transmission** (as described by study authors): ‘droplet infection and personal contact with the other victims’. |
| **Plague diagnosis** | **Definitions of plague cases:** not reported  **Laboratory findings:** ‘The first three or four of this series were thought to be influenza with pneumonia’ and did not have microbiological diagnosis as plague was not suspected yet.  Autopsy of case J: lung slides showed ‘numerous bipolar staining bacilli resembling plague’. Autopsy of case M: bacterial identification of plague (culture from guinea pig inoculated with lung tissue).  Sputum samples of case H have been reported with identification of pneumococci. |
| **Contacts** | **Number of contacts:** not reported  **Attack rate**: not reported  **Method of contact tracing:** not reported  **Definition of ‘contact’:** not reported  **Other relevant data describing contacts:** none |
| **Other relevant notes** | None |

| **Quality appraisal** | **Authors’ judgment** | **Support for judgment** |
| --- | --- | --- |
| 1. Were patient characteristics adequately reported? | Yes | Description of index cases and infected contacts were adequately reported. |
| 2. Was there some effort to trace all contacts from the index case? | Unknown | Not reported |
| 3. Were the methods used for tracing contacts adequate? | Not applicable | Not applicable |
| 4. Were the laboratory methods used for defining a confirmed case of plague reliable? | Yes | Initially, plague was not suspected. After suspicion of plague, patients were isolated and the outbreak was controlled.  The diagnosis was confirmed with laboratory methods in one case by a positive culture for *Y. pestis* from guinea pig inoculated with lung tissue of the infected case.  The autopsy of ‘one of the other cases of the series’ included smears from lung and spleen that showed numerous typical bipolar staining organisms. Although microbiological confirmation was not performed for the other cases, the clinical and epidemiological context together with microbiological confirmation of plague in one case is highly suggestive of an outbreak of pneumonic plague. |
| 5. Was the route of transmission plausible? | Yes | By respiratory route |
| 6. Was the cause-effect of transmission plausible? | Yes | Context of pneumonic plague outbreak following the route of transmission described above, from an index case with secondary pneumonic plague. |

| **Kugeler 2015** | |
| --- | --- |
| **Basic information** | **Setting:** United States  **Date:** 1900 to 2012 (patients infected through person-to-person route reported from 1900 to 1925)  This manuscript summarized the cases of plague in the US from 1900 to 2012, from the US Public Health Service, the Centers for Disease Control and Prevention (CDC), state reports and peer-reviewed publications. We report here the cases of plague from this manuscript associated with person-to-person transmission only. |
| **Index cases** | **Number of index cases:** not reported  **Form of plague:** not reported  **Description of infected cases:** not reported |
| **Infected cases from index cases** | **Number of contacts infected:** 49  **Form of plague:** pneumonic plague  **Description of infected contacts:** no disaggregated data among the 496 cases of plague during this period of time.  **Route of transmission** (as described by study authors): not reported |
| **Plague diagnosis** | **Definitions of plague cases: ‘**clinically compatible human illness and at least 1 of the following: 1) *Y. pestis* isolated from or detected in a clinical specimen, 2) elevated antibody titer to *Y. pestis* F1 antigen in >1 serum specimen, or 3) supportive epidemiologic and other laboratory evidence (e.g., visualization of typical *Y. pestis* morphology on a stained slide).’  ‘The clinical form of plague (e.g., bubonic, pneumonic, septicemic) was determined on the basis of explicit notations in the case records or from available clinical details; only the primary clinical form was considered. For example, patients who had primary bubonic plague and secondary pneumonic plague were classified as having bubonic plague.’  **Laboratory findings:** not reported |
| **Contacts** | **Number of contacts:** not reported  **Attack rate**: not reported  **Method of contact tracing:** not reported  **Definition of ‘contact’:** not reported  **Other relevant data describing contacts:** none |
| **Other relevant notes** | None |

| **Quality appraisal** | **Authors’ judgment** | **Support for judgment** |
| --- | --- | --- |
| 1. Were patient characteristics adequately reported? | Partial | Description of the plague cases are well reported for the overall cases reported in the manuscript, but with no disaggregated data for the cases of plague attributed to human to human transmission. |
| 2. Was there some effort to trace all contacts from the index case? | Unknown | Not reported. |
| 3. Were the methods used for tracing contacts adequate? | Not applicable | Not applicable |
| 4. Were the laboratory methods used for defining a confirmed case of plague reliable? | Yes | Clear definitions of plague are provided |
| 5. Was the route of transmission plausible? | Unknown | The route of infection is described to be ‘person-to-person’ without providing further information. |
| 6. Was the cause-effect of transmission plausible? | Unknown | Same as above |

| **Rabaan 2019** | |
| --- | --- |
| **Basic information** | **Setting:** Madagascar, in a non-endemic area and in large urban centers, including the capital city Antananarivo.  **Date:** August to November 2017 |
| **Index cases** | **Number of index cases:** 1  **Form of plague:** not fully detailed, but developed respiratory symptoms, so probably secondary pneumonic plague.  **Description of infected cases:** ‘a 31-year-old man from Toamasina who developed malaria-like symptoms’. He developed respiratory symptoms 4 days later with a fatal outcome. Respiratory symptoms when travelling in a public taxi. |
| **Infected cases from index cases** | **Number of contacts infected:** 31 contacts from the single index case. 2417 cases in total during the outbreak.  **Form of plague:** pneumonic plague  **Description of infected contacts:** not reported.  **Route of transmission** (as described by study authors): ‘ready transmission by airborne droplets’ ‘Pneumonic transmission occurs person-to-person via respiratory droplets, facilitated by the densely populated nature of the urban centres.’ |
| **Plague diagnosis** | **Definitions of plague cases:** following WHO definitions of confirmed, probable and suspected cases of plague.  **Laboratory findings:** not reported at an individual level. However, laboratory methods would include diagnostic tests allowing classification of cases as per the WHO definitions: isolation of *Y pestis*, serology, immunochromatography, and PCR. |
| **Contacts** | **Number of contacts:** not reported.  **Attack rate**: not reported.  **Method of contact tracing:** not reported. Active surveillance established in Madagascar.  **Definition of ‘contact’:** not reported  **Other relevant data describing contacts:** none |
| **Other relevant notes** | ‘For the index case, initially there was no suspicion of plague and so his body was prepared for burial using traditional methods, without any special precautions. Funerary practices have been previously observed to coincide with plague onset in Madagascar, in particular spread of pneumonic plague’  ‘Pneumonic plague patients should be isolated, masks should be provided for both patients and HCWs to reduce droplet transmission, bedding, clothing, sputum and excreta should be treated with chlorinated solution, and infection prevention and control measures should be observed by HCWs.’  ‘The current plague outbreak in Madagascar highlights the rise in importance of pneumonic plague and how its transmission from person to person can have devastating impacts in the context of overcrowded urban communities.’ |

| **Quality appraisal** | **Authors’ judgment** | **Support for judgment** |
| --- | --- | --- |
| 1. Were patient characteristics adequately reported? | Partial | Some data are provided for the case index, but very limited characteristics are reported regarding the infected contacts. The number of cases with pneumonic plague was found from another manuscript. |
| 2. Was there some effort to trace all contacts from the index case? | Partial | Active surveillance from Madagascar is assumed to happen but not clearly reported. |
| 3. Were the methods used for tracing contacts adequate? | Unknown | Not reported. |
| 4. Were the laboratory methods used for defining a confirmed case of plague reliable? | Yes | Cases were defined as confirmed, probable and suspected cases of plague according to the WHO definitions. |
| 5. Was the route of transmission plausible? | Yes | Authors attributed the transmission ‘via respiratory droplets’ |
| 6. Was the cause-effect of transmission plausible? | Yes | Context of pneumonic plague outbreak following the route of transmission described above. |

| **Ramasindrazana 2017** | |
| --- | --- |
| **Basic information** | **Setting:** Madagascar, in a remote area that had been free of human plague for 13 years.  **Date:** August 2015 |
| **Index cases** | **Number of index cases:** 1 (case A)  **Form of plague:** the authors suggest that case index (A) had bubonic plague from rodents or fleas, which developed to secondary pneumonic plague.  **Description of infected cases:** 22-year-old man, who developed chest pain, fever and cough 1 week after returning home from travelling. He died and ’was buried in a traditional manner with a 2-night wake, exposing the family and community to the pathogen and initiating a chain of transmission.’ |
| **Infected cases from index cases** | **Number of contacts infected:** from case index A, 11 cases (2 from immediate family, 6 from extended family, 3 from the community). Two additional cases were infected from these above cases.  **Form of plague:** pneumonic plague  **Description of infected contacts:** 9 man and 4 women, median age 22.5 years (range from 15 to 80). All of them had cough, 93% of them had blood-stained sputum.  **Route of transmission** (as described by study authors):  ‘The matched genetic grouping between the 2 human samples is consistent with human-to-human transmission.’ |
| **Plague diagnosis** | **Definitions of plague cases:** ‘according to the international standards definitions’  **Laboratory findings:** 4 cases of confirmed plague (2 by culture, 2 by seroconversion), 1 presumptive, 9 suspected (as no samples collected from these 9 cases with fatal outcome) |
| **Contacts** | **Number of contacts:** 123 from the index case (A)  **Attack rate**: not reported. But reproductive number (R_0_): 1.44 and transmission rate: 0.41 susceptibles/day.  **Method of contact tracing:** outbreak investigation protocol established by the Institut de Pasteur de Madagascar and the Malagasy Ministry of Health.  **Definition of ‘contact’:** not reported  **Other relevant data describing contacts:** post-exposure prophylaxis was given to the 35 contacts with positive serology. |
| **Other relevant notes** | ‘During pneumonic plague outbreaks, person-to-person transmission facilitates the spread from the initial infected person to family members and the wider community.’  ‘Pneumonic plague is rare but persists as a threat in Madagascar, where poor healthcare systems and traditional burial practices promote these outbreaks.’ |

| **Quality appraisal** | **Authors’ judgment** | **Support for judgment** |
| --- | --- | --- |
| 1. Were patient characteristics adequately reported? | Yes | Description of index cases and infected contacts were adequately reported. |
| 2. Was there some effort to trace all contacts from the index case? | Yes | Contacts were traced and reported. |
| 3. Were the methods used for tracing contacts adequate? | Yes | Following the outbreak investigation protocol established by the Institut de Pasteur de Madagascar and the Malagasy Ministry of Health. |
| 4. Were the laboratory methods used for defining a confirmed case of plague reliable? | Yes | Cases were defined according to ‘international standard definitions’ and classified as confirmed, presumptive and suspected, with laboratory confirmation detailed. |
| 5. Was the route of transmission plausible? | Yes | Although not directly stated by the authors, it is assumed from the manuscript to be via respiratory transmission. |
| 6. Was the cause-effect of transmission plausible? | Yes | Index case with respiratory symptoms and exposed to the family and community as he was ‘buried in a traditional manner with a 2-night wake’. |

| **Ratsitorahina 2000** | |
| --- | --- |
| **Basic information** | **Setting:** Madagascar, in a remote village of the central highlands  **Date:** October and November 1997 |
| **Index cases** | **Number of index cases:** 1  **Form of plague:** suspected bubonic plague with secondary pneumonic plague  **Description of infected cases:** woodcutter who developed fever and tender axillary adenitis. He later developed chest pain, blood-stained sputum and cough. Fatal outcome. |
| **Infected cases from index cases** | **Number of contacts infected:** 17 in total, from index case and among them.  **Form of plague:** primary pneumonic plague  **Description of infected contacts:** including the index case, nine males and nine females, with median age of 37 years including two children.  The healer who has been in direct contact with the index case, ‘incised the patient’s epigastric region and sucked out some blood’  ‘Developed severe fever, dyspnea, chest pain, diarrhea, and coughing with foamed and bloody sputum.’  All cases were from close contact with the index case or among them: the healer of the index case, healer’s family and patient, and villagers who had stayed in the healer’s house for the funeral ceremony. Other patients who developed plague attended the healer’s funeral or nursed plague patients.  **Route of transmission** (as described by study authors): ‘the contamination between patients is due to the direct transmission of *Yersinia pestis* through infective cough droplets’ |
| **Plague diagnosis** | **Definitions of plague cases:** not reported  **Laboratory findings:** by culture, immunochromatography and direct F1 ELISA from sputum samples and serology. No samples from the first 5 cases as buried before plague was detected. Only one patient was negative for all the tests done and it was concluded that he did not have plague. |
| **Contacts** | **Number of contacts:** 154 (not including those who get infected)  **Infection rate**: 8.4%  **Method of contact tracing:** not reported other than national active surveillance  **Definition of ‘contact’:** not reported  **Other relevant data describing contacts:** post-exposure chemoprophylaxis was given to the identified contacts. |
| **Other relevant notes** | ‘The patients […]. Their infection resulted from their active participation in the funeral ceremonies and attendance on patients. Patients with pneumonic plague are known to be contagious at the end-stage of the disease and the number of passages of *Y pestis* in human lungs seems to increase its virulence.’  ‘The risk of spreading pneumonic plague is actually not as high as may be thought. By the use of IgG anti-F1 ELISA, of which the specificity was 98·5% in Madagascar, we were able to estimate the infection rate in the contact population as 8·4%. The chance of a previous exposure to *Y pestis* is negligible since human plague has not been seen in these villages for 50 years.’  ‘Elementary hygiene measures to protect family members or health workers, such as the isolation of the patient and wearing of a mask, easily prevent contagion.’ |

| **Quality appraisal** | **Authors’ judgment** | **Support for judgment** |
| --- | --- | --- |
| 1. Were patient characteristics adequately reported? | Yes | Description of index cases and infected contacts were adequately reported. |
| 2. Was there some effort to trace all contacts from the index case? | Yes | Contacts were traced and reported. |
| 3. Were the methods used for tracing contacts adequate? | Unknown | Not reported |
| 4. Were the laboratory methods used for defining a confirmed case of plague reliable? | Partial | Some cases were retrospectively diagnosed with no laboratory diagnosis as already buried; efforts were made to confirm cases of plague in other patients by reliable methods. |
| 5. Was the route of transmission plausible? | Yes | Inhalation of ‘infective cough droplets’ |
| 6. Was the cause-effect of transmission plausible? | Yes | The index case presented ‘blood-stained sputum and cough’, and the infected contact developed primary pneumonic plague. |

| **Richard 2015** | |
| --- | --- |
| **Basic information** | **Setting:** Madagascar, in a Northern remote region that was supposedly free of *Y pestis*.  **Date:** 2011 |
| **Index cases** | **Number of index cases:** 1 (index case A)  **Form of plague:** primary vs secondary pneumonic plague (no bubo described)  **Description of infected cases:** 13-year-old boy working in a copper mine, who developed fever, headache and chills in the journey back home (50km distance). He later developed severe chest pain, cough and hemoptysis. Fatal outcome 8 days after onset of symptoms. |
| **Infected cases from index cases** | **Number of contacts infected:** 4 cases (B) infected from index case (A) + 15 cases from contact with Cases (B). Overall, 19 cases of plague by human-to-human transmission.  **Form of plague:** pneumonic plague  **Description of infected contacts:** close contacts including family members and care takers. All patients had sudden onset of fever, cough, hemoptysis and chest pain.  **Route of transmission** (as described by study authors): not directly stated, but authors mention in the introduction that ‘If the pathogen is transmitted as an aerosol by droplets or by contaminated dust, primary pneumonic plague may result.’ In the context of a human outbreak. |
| **Plague diagnosis** | **Definitions of plague cases:** according to WHO definitions.  **Laboratory findings:** limited samples collected as no post-mortem samples and plague declared after death of several cases. Subsequently to culture, serology, immunochromatography and molecular analysis: 17 suspected cases, 3 confirmed cases (and 2 presumptive cases from contacts who presented positive serology). |
| **Contacts** | **Number of contacts:** 41 (not including infected cases). 39 of them received chemoprophylaxis. Health personnel also received chemoprophylaxis.  **Attack rate**: 55%  **Method of contact tracing:** not reported  **Definition of ‘contact’:** ‘have interact with the patients’. ‘Family contacts: persons who lived in the same household as an infected person during the outbreak’, including single-room houses.  **Other relevant data describing contacts:**  Some contacts were people who ‘had spent some time with a patient or approached a patient who died during the outbreak’, others were direct family members living in the same house. One contact shared the same bed with a plague case until his death and was not infected. Ten contacts ‘had attended funerals for case-patients in different villages’ |
| **Other relevant notes** | ‘At this lethal stage of the disease, which lasts ≤3 days, patients are highly infectious.’  ‘During the latency period before hemoptysis, sputum contains hardly any infectious organisms. Simple countermeasures, such as protective facial masks, are efficient in preventing transmission by droplets. Also, turning one’s head away from or turning one’s back to- ward a healthy person has a major prophylactic effect.’  ‘It has been suggested that patients with bubonic plague and patients who have died of plague are not directly infectious to other humans’ ‘This suggestion is consistent with findings in the present study because contacts [10 of them] who only attended the funerals did not show symptoms or seroconversion’ |

| **Quality appraisal** | **Authors’ judgment** | **Support for judgment** |
| --- | --- | --- |
| 1. Were patient characteristics adequately reported? | Yes | Description of index cases and infected contacts were adequately reported. |
| 2. Was there some effort to trace all contacts from the index case? | Yes | Contacts were traced and reported. |
| 3. Were the methods used for tracing contacts adequate? | Unknown | Not reported. |
| 4. Were the laboratory methods used for defining a confirmed case of plague reliable? | Yes | Cases were defined as confirmed, probable and suspected cases of plague according to the WHO definitions. |
| 5. Was the route of transmission plausible? | Yes | Although not directly stated by the authors, it is assumed from the manuscript to be ‘transmitted as an aerosol by droplets or by contaminated dust’. |
| 6. Was the cause-effect of transmission plausible? | Yes | Index case and infected contacts with pneumonic plague |

**S6 Appendix. Summary of studies describing plague acquired from human and animal cadavers (review part 2)**

| **Study ID** | **Study design**  **Setting, date** | **Source of infection** | **Cases infected**  **(form of plague*)** | **Time from animal death to exposure** | **Description of exposure** | **Route of transmission according to study authors** |
| --- | --- | --- | --- | --- | --- | --- |
| CDC 1992 (29) | Case series  US, 1992 | Ground squirrel | 1^†^ (BP) | Unknown | Skinned and consumed | Not reported |
| Christie 1980 (36) | Case series  Libya, 1976 | Camel | 12^‡^ (7 BP axillary, cervical) | 4 immediate  8 unknown | 4 slaughtered and skinned the camel  1 distributed the meat  7 ate or handled the camel meat | Direct handling  Consumption of camel meat |
|  |  | Goat | 5 (form not reported) | 1 immediate  4 unknown | 1 killed and skinned the goat  1 treated the skin  3 same household where skin was kept | Direct handling |
| Gage 2000 (30) | Case series  US, 1984 | Domestic cat | 1 (BP, axillary) | Unknown | Buried the dead cat | Direct contact from infectious body fluids of the cat cadaver |
| Ge 2015 (26) | Case series  China, 2000-12 | Fox, marmots, dogs | 32 (25 PP, 7 BP) | Unknown | 18 flayed  12 buried  1 gave to feed a dog  1 captured by a dog | Not reported |
|  | Case report  China, 2014 | Marmot | 1 (PP) | Unknown | Seized from a dog | Exposure to aerosols |
| Kartman 1960 (32) | Case series  US, 1908-60 | Wild rabbits  (cottontail rabbits) | 5 (form not reported) | Unknown | Killed and cleaned diseased animal cadavers | Direct handling |
|  |  |  | 4 (BP, axillary) | Unknown | 2 handled and skinned 6 to 8 rabbits  2 shot and skinned 8 or 9 rabbits with bare hands ‘which became contaminated with blood, body fluids and bits of tissue.’ ‘The hands of both men had been cut and abraded by mesquite thorns.’ | Direct handling |
| Kartman 1970 (31) | Case series  US, 1908-68 | Ground squirrel | 1 (BP) | Unknown | Hunted the animal | Direct handling |
|  |  | Ground squirrels, rabbits, prairie dogs, kangaroo rat, pocket gophers | 16^§^ (BP) | Unknown | Contact with infected animals:  12 shot/killed  1 handled  1 cut himself on a rabbit bone  1 unsterile autopsy  1 played | Direct handling |
|  |  | Prairie dog | 1 (form not reported) | Unknown | Hunted the animal | Direct handling |
| Kugeler 2015^¶^ (33) | Synopsis  US, 1900-2012 | Animals | 64 (58 BP) | Unknown | Butchered or skinned an animal | Not reported |
| Mitchell 1930 (38) | Report  South Africa, 1930 | Human cadavers | 1 (BP, axillary) | Unknown | Postmortem examination of 2 human cadavers | Not reported |
| Poland 1973 (34) | Case report  US, 1972 | Bobcat (*Lynx rufus*) | 1 (BP, epitrochlear) | Immediate and within <24h | Animal was shot during the day and put on the vehicle. In the evening, the person who got infected (had open lesions on his hands) hold the animal with another student while a third student eviscerated and skinned the animal. The 2 other students had no known open lesions on their hand or arms and were not infected. | Direct contact through breaks in the skin. |
| Ratsito-rahina 2000 (22) | Case series  Madagascar, 1997 | Human cadavers | 9 (PP) | Probably around <24-36h | 8 stayed for 2 days at the healer’s house for funeral ceremony of the healer who died from plague. This coincides with the last 2 days of life of the healer’s wife and son who also died from plague. Therefore, exposure to human cadaver but also to live humans with the disease.  1 man who attended the healer’s funeral. Similarly, possibility of interhuman transmission.  Authors mentioned ‘other villagers became infected during the funeral ceremonies’ | ‘Infection resulted from active participation in the funeral ceremonies and attendance on patients.’ |
| Saeed 2005 (39) | Case series  Saudi Arabia, 1994 | Camel | 1^#^ (BP, axillary) | Within <24h | Slaughtered and cut his arm while killing the animal | Not reported  (Probably direct contact through break in the skin) |
| Sagiev 2019 (37) | Case series  Kazakhstan, 1974-2003 | Camel, hare, saiga | 12 (form not reported) | Unknown | 8 slaughter of camel  2 cutting carcass of hare  1 handling corpse of hare  1 cutting a sick saiga | Not reported |
| Von Reyn 1976 (35) | Case report  US, 1975 | Coyote | 1 (BP, axillary) | Unknown | Skinned the animal and carried the pelt  Had forearm laceration and nailbed exposed during skinning | Direct contact through break in the skin |
| Wong 2009 (8) | Case report  US, 2007 | Mountain lion | 1 (PP) | Around 35h | Carried the carcass for approximately 1 km to his vehicle, then into his garage.  Performed necropsy with bare hands, including the opening of the animal’s thoracic cavity, found filled with blood, and transecting the vertebral column. Estimated time of exposure during the necropsy: 2.5 hours. | Inhalation of aerosols generated while handling the infected animal. |
| Wu 2009 (27) | Case series  China, 1975-2007 | Tibetan sheep | 25 (9 BP, 6 PP, 3 SP, 2 intestinal plague) | Unknown | Flayed, ate or handled animal cadavers | Not reported |
| Zhang 2007 (28) | Case series  China, 1958-2005 | Marmot, cat and human cadavers | 56 (28 PP, 21 BP, 5 SP^**^) | Unknown | 24 flayed, ate or handled marmots or cats (live or cadavers); NDD  32 infected by contacting with plague patients or human cadaver; NDD | Not reported |

Abbreviations: BP: bubonic plague; CDC: Centers for Disease Control and Prevention; NDD: no disaggregated data; PP: pneumonic plague; SP: septicemic plague; US: the United States.

^*^We report forms of primary plague and do not detail development of secondary forms of plague.

^†^Other cases of plague are reported but related with flea or unknown transmission.

^‡^Three children (children of meat dealer) were also reported in this case report. However, 2 of them were sick before the contact with the camel and were diagnosed as typhoid fever, and the third one might be infected with plague but had no direct contact with the camel.

^§^Authors reported that ‘specific animal contact is known to have occurred a few days prior to illness in 35 of the 80 bubonic cases’. Among these 35 cases, we hereby only reported the 16 cases in which transmission was described from a dead animal and did not report those infected from contact with alive animals (such as bites) or unclear cases.

^¶^ This is a synopsis that reports the cases of plague in the US from 1900 to 2012. Route of exposure was documented for 30% of the cases. We report here those attributed to animal cadavers (butchering or skinning) and exclude cases for which transmission from animal cadaver was unclear such as animal handling.

^#^ Four other cases of plague (pharyngeal plague) are described in this manuscript, who ate raw meat of the camel. The case we report in this table had not eaten camel meat.

^**^These numbers of different forms of plague are among the total of 64 cases reported in the manuscript, including 8 cases of unknown route of transmission, that we excluded for the purpose of this review.

**S7 Appendix. Characteristics of included studies for plague acquired from human and animal cadavers (review part 2)**

| **CDC 1992** | |
| --- | --- |
| **Setting** | **Country:** the United States  **Study dates:** April 1992 |
| **Source of infection** | **Source of infection:** Ground squirrel (*Spermophilus beldingi*)  **Plague diagnosis:** not reported  **Other possible sources of infection:** not reported |
| **Exposure to the source of infection** | **Description of exposure:** skinned the animal cadaver and consumed the meat  **Duration of exposure:** not reported  **Time between death of the human/animal and contact with the infected human:** not reported |
| **Persons infected** | **Number:** 1  **Age, sex:** 16 years, male  **Profession:** not reported  **Signs and symptoms:** not reported  **Form of plague:** bubonic and secondary septicaemic plague  **Plague diagnosis:** positive blood culture  **Outcome:** not reported |
| **Route of transmission** | **Attributed route of transmission by the study author:** not reported  **Plausibility of the cause-effect transmission as per the study authors:** not reported |
| **Other relevant notes** | **Cases exposed to the same source but not infected:** not reported |

| **Quality appraisal** | **Authors’ judgment** | **Support for judgment** |
| --- | --- | --- |
| 1. Were patient characteristics adequately reported? | Yes | Although more details could have been provided, adequate description of the infected case |
| 2. Was there some effort to trace all contacts from the index case? | Unknown | Not reported |
| 3. Were the methods used for tracing contacts adequate? | Not applicable | Not applicable |
| 4. Were the laboratory methods used for defining a confirmed case of plague reliable? | Yes | Positive blood culture |
| 5. Was the route of transmission plausible? | Yes | Not specifically describe by the study authors, but transmission through direct contact plausible from the infected cadaver through the skin of the person while skinning, with subsequent bubonic plague. |
| 6. Was the cause-effect of transmission plausible? | Yes | As above |

| **Christie 1980** | | |
| --- | --- | --- |
|  | **Case series 1** | **Case series 2** |
| **Setting** | **Country:** North East Libya  **Study dates:** February 1976 | **Country:** North East Libya  **Study dates:** June 1976 |
| **Source of infection** | **Source of infection:** camel  **Plague diagnosis:** not reported  **Other possible sources of infection:** none reported | **Source of infection:** goat  **Plague diagnosis:** not reported  **Other possible sources of infection:** 4 dead rats found in the compound |
| **Exposure to the source of infection** | **Description of exposure:**  A) 4 people: slaughtered the camel  B) 1 person: distributed the meat  C) 7 people: handled or eaten camel meat  **Duration of exposure:** not reported  **Time between death of the human/animal and contact with the infected human:**  A) before and immediately after killing  B and C) not specified | **Description of exposure:**  A) 1 person: killed and skinned the goat  B) 1 person: treated the skin  C) 3 people: no direct exposure, from the same house  **Duration of exposure:** not reported  **Time between death of the human/animal and contact with the infected human:** A) immediately,  B and C) not reported |
| **Persons infected** | **Number:** 12  **Age, sex:** not reported  **Profession:** B) meat dealer, others not reported  **Signs and symptoms:** A and B) not reported; C) axillary or neck buboes  **Form of plague:** A and B) not reported, C) bubonic plague.  **Plague diagnosis:** A and B) could not undergo laboratory diagnosis  C) 7 diagnosed by hemagglutination titer  **Outcome:** A) and B) death C) recovery | **Number:** 5  **Age, sex:** A) adult male; B) adult female; C) adult female, two children  **Profession:** not reported  **Signs and symptoms:** not reported  **Plague diagnosis:** serology positive in four, method not reported  **Outcome:** recovery |
| **Route of transmission** | **Attributed route of transmission by the study author:** direct handling and/or eating camel meat  **Plausibility of the cause-effect transmission as per the study authors:** plausible | **Attributed route of transmission by the study author:** direct contact  **Plausibility of the cause-effect transmission as per the study authors:** not clearly reported |
| **Other relevant notes** | **Cases exposed to the same source but not infected:** unknown number of villagers ate meat. | **Cases exposed to the same source but not infected:** not reported |

| **Quality appraisal** | **Authors’**  **judgment (Case series 1)** | **Support for judgment** | **Authors’**  **judgment (Case series 2)** | **Support for judgment** |
| --- | --- | --- | --- | --- |
| 1. Were patient characteristics adequately reported? | Partial | Some of the patient characteristics not described | Partial | Some of the patient characteristics not described |
| 2. Was there some effort to trace all contacts from the index case? | Unknown | No details on other contacts | Unknown | No details on other contacts |
| 3. Were the methods used for tracing contacts adequate? | Not applicable | Not applicable | Not applicable | Not applicable |
| 4. Were the laboratory methods used for defining a confirmed case of plague reliable? | Partial | Laboratory diagnosis made in 7 cases | Partial | Serology performed in 4 cases |
| 5. Was the route of transmission plausible? | Yes | All infected persons had close contact with the infected animal, by either direct handling or consumption of the meat | Partial | Yes, by direct contact in 2 cases. However, unclear for the 3 cases who did not have direct contact with the infected animal but were sharing the household with the infected humans. |
| 6. Was the cause-effect of transmission plausible? | Yes | As above | Partial | As above |

| **Gage 2000** | |
| --- | --- |
| **Setting** | **Country:** California  **Study dates:** March 1984 |
| **Source of infection** | **Source of infection:** cat  **Plague diagnosis:** clinically (laboratory confirmation not done)  **Other possible sources of infection:** inactive rodent burrows suggestive of epizootic were noted at the site (reaffirming plague as a cause of infection for the cat) |
| **Exposure to the source of infection** | **Description of exposure:** buried a dead cat  **Duration of exposure:** not reported  **Time between death of the human/animal and contact with the infected human:** not reported |
| **Persons infected** | **Number:** 1  **Age, sex:** 24 years, male  **Profession:** not reported  **Signs and symptoms:** cellulitis, axillary bubo, thrombocytopenia, gastrointestinal bleeding, acute respiratory distress syndrome, lactic acidosis  **Form of plague:** bubonic plague  **Plague diagnosis:** positive bacterial culture  **Outcome:** death |
| **Route of transmission** | **Attributed route of transmission by the study author:** direct contact from infectious body fluids of the cat cadaver; entry route not specified  **Plausibility of the cause-effect transmission as per the study authors:** plausible, as above |
| **Other relevant notes** | **Cases exposed to the same source but not infected:** none reported |

| **Quality appraisal** | **Authors’ judgment** | **Support for judgment** |
| --- | --- | --- |
| 1. Were patient characteristics adequately reported? | Yes | Relevant patient characteristics described |
| 2. Was there some effort to trace all contacts from the index case? | Unknown | No information on contacts |
| 3. Were the methods used for tracing contacts adequate? | Not applicable | Not applicable |
| 4. Were the laboratory methods used for defining a confirmed case of plague reliable? | Yes | Diagnosed using isolation of the organism |
| 5. Was the route of transmission plausible? | Yes | Direct contact through infected fluids |
| 6. Was the cause-effect of transmission plausible? | Yes | Axillary bubo as a consequence of handling an infected cadaver is plausible |

| **Ge 2015** | | |
| --- | --- | --- |
|  | **Case report** | **Case series** |
| **Setting** | **Country:** China  **Study dates:** July 2014 | **Country:** China  **Study dates:** 2000 to 2012 |
| **Source of infection** | **Source of infection:** marmot  **Plague diagnosis:** not assessed, but five dogs fed with the marmot were F1 antigen positive.  **Other possible sources of infection:** not reported | **Source of infection:** fox, marmots, dog  **Plague diagnosis:** not reported  **Other possible sources of infection:** not reported |
| **Exposure to the source of infection** | **Description of exposure:** seized from a dog  **Duration of exposure:** short period  **Time between death of the human/animal and contact with the infected human:** not reported, likely immediate | **Description of exposure:** 18 flayed, 12 buried, 1 gave to feed a dog, 1 captured by a dog  **Duration of exposure:** not reported  **Time between death of the human/animal and contact with the infected human:** not reported |
| **Persons infected** | **Number:** 1  **Age, sex:** 38 years, Male  **Profession:** shepard  **Signs and symptoms:** fever, bilateral lung signs, left pleural effusion, pericardial effusion, dilated intestines, shock  **Form of plague:** primary pneumonic plague  **Plague diagnosis:** reverse IHA for F1 antigen positive in serum (1:40), throat (1:6400) and sputum (1:12800). PCR positive. Culture positive from sputum, throat swab and blood.  **Outcome:** death | **Number:** 32  **Age, sex:** not reported  **Profession:** not reported  **Signs and symptoms:** not reported  **Form of plague:** 25 pneumonic plague;  7 bubonic plague  **Plague diagnosis:** not reported **Outcome:** no disaggregated data |
| **Route of transmission** | **Attributed route of transmission by the study author:** exposure to aerosols  **Plausibility of the cause-effect transmission as per the study authors:**  likely with primary pneumonic plague | **Attributed route of transmission by the study author:** not reported  **Plausibility of the cause-effect transmission as per the study authors:** not reported |
| **Other relevant notes** | **Cases exposed to the same source but not infected:** brother of the patient dismembered the cadaver and fed the dogs | **Cases exposed to the same source but not infected:** not reported |

| **Quality appraisal** | **Authors’**  **judgment (Case report)** | **Support for judgment** | **Authors’**  **judgment (Case series)** | **Support for judgment** |
| --- | --- | --- | --- | --- |
| 1. Were patient characteristics adequately reported? | Yes | All patient characteristics described in detail | Partial | Some of the patient characteristics not described |
| 2. Was there some effort to trace all contacts from the index case? | Yes | Contacts who were not infected were described in detail | Unknown | Not described |
| 3. Were the methods used for tracing contacts adequate? | Yes | Well described, with paired serology performed to all contacts | Not applicable | Not applicable |
| 4. Were the laboratory methods used for defining a confirmed case of plague reliable? | Yes | Several validated laboratory methods | Unknown | Not reported |
| 5. Was the route of transmission plausible? | Yes | Transmission via aerosols described | Partial | Route of transmission not described. Mostly pneumonic plague due to inhalation of aerosols generated while handling the infected cadavers? |
| 6. Was the cause-effect of transmission plausible? | Yes | Primary pneumonic plague due to aerosol transmission | Partial | As above |

| **Kartman 1960** | |
| --- | --- |
| **Setting** | **Country:** the United States (California and New Mexico)  **Study dates:** from 1908 until 1960 |
| **Source of infection** | **Source of infection:** wild rabbits (cottontail rabbits)  **Plague diagnosis:** not reported  For 1 case, authors specified that domestic and wild mammals were collected in the area for investigation and prove presence of plague. All 4 animals found dead were infected with plague, and 2 were cottontail rabbits.  **Other possible sources of infection:**  For 2 cases: no evidence of flea bites, and investigation in the area on wild animals and fleas showed infected animals with plague. |
| **Exposure to the source of infection** | **Description of exposure:**  For 5 cases: “The California infections were acquired after the victims killed and cleaned brush rabbits.”  For 1 case: “The victim became ill 3 days after he had skinned 6 cottontail rabbits shot near Maljamar”  For 1 case: “The patient shot and dressed 8 cottontail rabbits and became ill with plague 4 days after”  For 2 cases: “They had hunted rabbits” and were hospitalized 4 and 6 days after. They had shot and skinned one rabbit on the spot the first night, and 8 or 9 rabbits the following day, which were skinned and dressed at home. “They were dressed with bare hands which became contaminated with blood, body fluids, and bits of tissue. The evidence showed that the hands of both men had been cut and abraded by mesquite thorns, that one of them had pulled several rabbits out of burrows with his bare hands, and also had ‘cleaned’ his hands by rubbing them with soil.”  **Duration of exposure:** not reported  **Time between death of the human/animal and contact with the infected human:** not reported |
| **Persons infected** | **Number:** 9  **Age, sex:** 4 cases were male adults, not reported for 5 cases  **Profession:** not reported  **Signs and symptoms:** axillary buboes in 4 cases, not reported for 5 cases  **Form of plague:** bubonic plague in 4 cases, not reported for 5 cases  **Plague diagnosis:** clinical in 2 cases, not reported for 7 cases  **Outcome:** 1 fatal outcome, 2 recoveries, rest of cases not reported. |
| **Route of transmission** | **Attributed route of transmission by the study author:** direct handling of diseased animal cadavers  **Plausibility of the cause-effect transmission as per the study authors:**  For 2 cases, “The victims in both cases had axillary buboes, which are consistent with their histories of having handled and skinned wild rabbits”  For 2 cases, “The location of lymphadenopathy and the incubation period were consistent with entrance of the etiologic agent by manual contact with infected rabbits” |
| **Other relevant notes** | **Cases exposed to the same source but not infected:** none reported. |

| **Quality appraisal** | **Authors’ judgment** | **Support for judgment** |
| --- | --- | --- |
| 1. Were patient characteristics adequately reported? | Partial | Characteristics including form of plague are given for four cases but not reported for the other 5 cases. |
| 2. Was there some effort to trace all contacts from the index case? | Unknown | No description of contacts from the 5 reported cases. |
| 3. Were the methods used for tracing contacts adequate? | Not applicable | Not applicable |
| 4. Were the laboratory methods used for defining a confirmed case of plague reliable? | No | Clinical diagnosis, or no details on confirmed diagnosis. However, clear methodology on microbiology diagnosis of plague in animals is given. |
| 5. Was the route of transmission plausible? | Yes | Authors described direct handling of the infected animal cadavers. |
| 6. Was the cause-effect of transmission plausible? | Yes | Quotes from text: “The victims in both cases had axillary buboes, which are consistent with their histories of having handled and skinned wild rabbits” (2 cases); “The location of lymphadenopathy and the incubation period were consistent with entrance of the etiologic agent by manual contact with infected rabbits” (2 cases) |

| **Kartman 1970** | |
| --- | --- |
| **Setting** | **Country:** the United States  **Study dates:** 1908 to 1968 |
| **Source of infection** | **Source of infection:**  (A) ground squirrel  (B) ground squirrels, rabbits, prairie dogs, kangaroo rat, pocket gophers  (C) prairie dog  **Plague diagnosis:** not reported  **Other possible sources of infection:** not reported |
| **Exposure to the source of infection** | **Description of exposure:**  (A) developed plague 3 to 4 days after hunting ground squirrels  (B) “Specific animal contact is known to have occurred a few days prior to illness in 35 of the 80 bubonic cases.” 4 had shot ground squirrels, 2 for food […]. 5 killed rabbits for sport, 1 for food, a boy cut himself on a rabbit bone, and 1 handled a rabbit brought to the house by her dog. […]. 2 killed prairie dogs.  “A biologist, studying prairie dogs, became ill after performing an unsterile autopsy on a dead prairie dog.”  “1 child played with a dead kangaroo rat”  “1 man killed pocket gophers”  (C) “The victim had hunted prairie dog  **Duration of exposure:** not reported  **Time between death of the human/animal and contact with the infected human:** not reported |
| **Persons infected** | **Number:** 18 (more cases are reported in the document, but with no specification on whether the persons got infected from alive or dead animals).  **Age, sex:** A) male adult; others not reported  **Profession:** A) laborer; others not reported  **Signs and symptoms: Not described**  **Form of plague:** (A) bubonic plague with secondary plague pneumonia  (B) bubonic plague  **Plague diagnosis:** not reported  **Outcome:** not reported |
| **Route of transmission** | **Attributed route of transmission by the study author:** direct handling  **Plausibility of the cause-effect transmission as per the study authors:** plausible |
| **Other relevant notes** | **Cases exposed to the same source but not infected:** none reported. |

| **Quality appraisal** | **Authors’ judgment** | **Support for judgment** |
| --- | --- | --- |
| 1. Were patient characteristics adequately reported? | Partial | Some salient characteristics of patients are not described |
| 2. Was there some effort to trace all contacts from the index case? | Unknown | No reporting on contact of cases |
| 3. Were the methods used for tracing contacts adequate? | Not applicable | Not applicable |
| 4. Were the laboratory methods used for defining a confirmed case of plague reliable? | Unknown | Laboratory methods of diagnosing cases were not described |
| 5. Was the route of transmission plausible? | Yes | Direct contact by handling infected animals (including killing them, autopsy and cutting himself with a bone), resulting in bubonic plague |
| 6. Was the cause-effect of transmission plausible? | Yes | As above |

| **Kugeler 2015** | |
| --- | --- |
| **Setting** | **Country:** the United States  **Study dates:** 1900 to 2012 |
| **Source of infection** | **Source of infection:** animal cadaver  **Plague diagnosis:** not reported  **Other possible sources of infection:** not reported |
| **Exposure to the source of infection** | **Description of exposure:** butchering or skinning of an animal cadaver  **Duration of exposure:** not reported  **Time between death of the human/animal and contact with the infected human:** not reported |
| **Persons infected** | **Number:** 64  **Age, sex:** not reported  **Profession:** not reported  **Signs and symptoms:** not reported  **Form of plague:** bubonic plague in 91% of cases  **Plague diagnosis:** not reported  **Outcome:** not reported |
| **Route of transmission** | **Attributed route of transmission by the study author:** not reported  **Plausibility of the cause-effect transmission as per the study authors:** not applicable |
| **Other relevant notes** | **Cases exposed to the same source but not infected:** Not described |

| **Quality appraisal** | **Authors’ judgment** | **Support for judgment** |
| --- | --- | --- |
| 1. Were patient characteristics adequately reported? | No | Patient characteristics not described |
| 2. Was there some effort to trace all contacts from the index case? | Unknown | Cannot assess from available information |
| 3. Were the methods used for tracing contacts adequate? | Not applicable | Not applicable |
| 4. Were the laboratory methods used for defining a confirmed case of plague reliable? | Unknown | Not reported |
| 5. Was the route of transmission plausible? | Partial | 64 cases are reported and attributed to butchering or skinning an animal. No further details are provided by the authors. Most of the cases were bubonic form of plague, with possible transmission through handling the animal cadaver. Other forms of plague are not described; pneumonic plague can result as inhalation of infected aerosols generated by butchering or skinning the cadaver. |
| 6. Was the cause-effect of transmission plausible? | Partial | As above. In addition, transmission via fleas might not have been fully excluded. |

| **Mitchell 1930** | |
| --- | --- |
| **Setting** | **Country:** South Africa  **Study dates:** November 1930 |
| **Source of infection** | **Source of infection:** 2 human cadavers  **Plague diagnosis:** confirmed by postmortem and laboratory diagnosis (test not specified)  **Other possible sources of infection:** field survey conducted in the area where it was indicated that plague was active. |
| **Exposure to the source of infection** | **Description of exposure:** performed postmortem examination of 2 cadavers  **Duration of exposure:** not reported  **Time between death of the human/animal and contact with the infected human:** not reported |
| **Persons infected** | **Number:** 1  **Age, sex:** adult male  **Profession:** district surgeon  **Signs and symptoms:** axillary buboes  **Form of plague:** bubonic plague  **Plague diagnosis:** not reported  **Outcome:** recovery |
| **Route of transmission** | **Attributed route of transmission by the study author:** not reported from the postmortem examination  **Plausibility of the cause-effect transmission as per the study authors:** plausible |
| **Other relevant notes** | **Cases exposed to the same source but not infected:** Not described |

| **Quality appraisal** | **Authors’ judgment** | **Support for judgment** |
| --- | --- | --- |
| 1. Were patient characteristics adequately reported? | Yes | Adequate description |
| 2. Was there some effort to trace all contacts from the index case? | Unknown | No details on other exposures |
| 3. Were the methods used for tracing contacts adequate? | Not applicable | Not applicable |
| 4. Were the laboratory methods used for defining a confirmed case of plague reliable? | Unknown | No description on which laboratory tests were used |
| 5. Was the route of transmission plausible? | Yes | Axillary bubonic plague due to handling infected bodies during the time involved for two autopsies is plausible |
| 6. Was the cause-effect of transmission plausible? | Yes | As above |

| **Poland 1973** | |
| --- | --- |
| **Setting** | **Country:** the United States (North Arizona)  **Study dates:** February 1972 |
| **Source of infection** | **Source of infection:** bobcat cadaver (*Lynx rufus*)  **Plague diagnosis:** *Y pestis* isolated from the bobcat brain tissue and bone marrow. Samples (brain and bone marrow) were taken 2 weeks after death.  **Other possible sources of infection:** field survey conducted in the area indicated that plague was active in the area. |
| **Exposure to the source of infection** | **Description of exposure:** dead animal was on the same vehicle during the day (inside, or in the boot, unspecified), then the patient hold the animal with another student, while a third student eviscerated and skinned the animal.  The student who skinned the animal “became extensively contaminated with blood and tissue contents from the animal”. The other 2 students (including the contaminated case) “were also contaminated but to a considerably lesser degree, since their primary task was to hold the animal [for the first student]”  “Following the skinning, the first student washed with soap and water; it was not ascertained how thoroughly the other 2 students (including the plague case) washed”  **Duration of exposure:** within the car (several hours); and the time of skinning the animal  **Time between death of the human/animal and contact with the infected human:** same day |
| **Persons infected** | **Number:** 1  **Age, sex:** 19 years, male  **Profession:** student  **Signs and symptoms:** onset with generalized myalgia, headache, pain in right elbow and shoulder, fever and upper respiratory symptoms.  Following day on admission: fever, anorexia, nausea, diarrhea.  During admission: chills, fever, anxiety, continued severe pain in right arm and shoulder. Epitrochlear and axillary lymphadenopathy  **Form of plague:** bubonic (epitrochlear) plague  **Plague diagnosis:** *Y pestis* identified from aspirate of the patient’s right epitrochlear lymph node. Paired sera (26 days apart): rise in titer for antibodies anti F1 of *Y pestis* from 1:4 to 1:32 by passive hemagglutination.  **Outcome:** recovery |
| **Route of transmission** | **Attributed route of transmission by the study author:** direct contact with contaminated animal through breaks in the skin.  **Plausibility of the cause-effect transmission as per the study authors:**  the 2 student who were presents but not contaminated “had no known open lesions on their hands or arms.” The contaminated student, however, “complained of ‘hang- nails’, was a ‘nail chewer’, and reported having numerous raw areas around his fingernails.” |
| **Other relevant notes** | **Cases exposed to the same source but not infected:** 2 other students exposed while they were skinning the bobcat, not infected; 2 other people with them during the day, not when skinning. |

| **Quality appraisal** | **Authors’ judgment** | **Support for judgment** |
| --- | --- | --- |
| 1. Were patient characteristics adequately reported? | Yes | All patient characteristics described in detail |
| 2. Was there some effort to trace all contacts from the index case? | Yes | Two contacts who were not infected were described in detail |
| 3. Were the methods used for tracing contacts adequate? | Yes | From death of the animal details described in sequence |
| 4. Were the laboratory methods used for defining a confirmed case of plague reliable? | Yes | Diagnosed using isolation of the organism |
| 5. Was the route of transmission plausible? | Yes | Direct contact with breaks of skin |
| 6. Was the cause-effect of transmission plausible? | Yes | With presence of epitrochlear nodes in bubonic plague route of transmission is likely |

| **Ratsitorahina 2000** | |
| --- | --- |
| **Setting** | **Country:** Madagascar  **Study dates:** October 1997 |
| **Source of infection** | **Source of infection:** human  **Plague diagnosis:** clinical  **Other possible sources of infection:** exposure to live humans with plague |
| **Exposure to the source of infection** | **Description of exposure:** 8 stayed for 2 days at the healer’s house for funeral ceremony of the healer who died from plague. 1 man who attended the healer’s funeral.  **Duration of exposure:** refer above  **Time between death of the human/animal and contact with the infected human:** probably around <24-36h (the healer was buried the day after he died) |
| **Persons infected** | **Number:** 9  **Age, sex:** 1 child (4 years), 8 adults (17- 65 years), 2 males, 7 females  **Profession:** not reported  **Signs and symptoms:** pneumonic syndrome  **Form of plague:** pneumonic plague  **Plague diagnosis:** rapid diagnostic test based on F1 antigen positivity  **Outcome:** recovery in 8, death in 1 |
| **Route of transmission** | **Attributed route of transmission by the study author:** ‘Infection resulted from active participation in the funeral ceremonies and attendance on patients’; ‘other villagers became infected during the funeral ceremonies.’  **Plausibility of the cause-effect transmission as per the study authors:** plausible |
| **Other relevant notes** | **Cases exposed to the same source but not infected:**  several tested, not clear about total number of exposed. 154 contacts tested. None infected, but 13 seropositive |

| **Quality appraisal** | **Authors’ judgment** | **Support for judgment** |
| --- | --- | --- |
| 1. Were patient characteristics adequately reported? | Yes | Adequate description of patient characteristics |
| 2. Was there some effort to trace all contacts from the index case? | Yes | Contacts were assessed clinically and serologically |
| 3. Were the methods used for tracing contacts adequate? | Unknown | Details not provided |
| 4. Were the laboratory methods used for defining a confirmed case of plague reliable? | Yes | Diagnosed using F1 rapid diagnostic test in sputum |
| 5. Was the route of transmission plausible? | Yes | Transmission of pneumonic plague via respiratory droplets |
| 6. Was the cause-effect of transmission plausible? | Partial | Pneumonic plague can be transmitted via respiratory droplets. However, it is unclear whether all the cases have been infected from human cadavers, as human to human transmission could be an alternative for some of them. Indeed, 8 stayed for 2 days at the healer’s house for funeral ceremony of the healer who died from plague. This coincides with the last 2 days of life of the healer’s wife and son who also died from plague. Similarly, possibility of interhuman transmission for one man described as attending the healer’s funeral. |

| **Saeed 2005** | |
| --- | --- |
| **Setting** | **Country:** Saudi Arabia  **Study dates:** February 1994 |
| **Source of infection** | **Source of infection:** camel  **Plague diagnosis:** *Y. pestis* isolated from bone marrow of camel and jirds and burrows  **Other possible sources of infection:** none |
| **Exposure to the source of infection** | **Description of exposure:** slaughtered and had a cut on his arm while killing  **Duration of exposure:** not reported  **Time between death of the human/animal and contact with the infected human:** same day |
| **Persons infected** | **Number:** 1  **Age, sex:** adult, male  **Profession:** not reported  **Signs and symptoms:** fever, axillary lymphadenitis and cellulitis  **Form of plague:** bubonic plague  **Plague diagnosis:** indirect hemagglutination positive  **Outcome:** recovery |
| **Route of transmission** | **Attributed route of transmission by the study author:** not clearly stated, but probably direct contact with contaminated animal through breaks in the skin.  **Plausibility of the cause-effect transmission as per the study authors:** plausible |
| **Other relevant notes** | **Cases exposed to the same source but not infected:** meat of the camel was distributed among 106 people, 37 people ate camel meat (only six people ate raw meat and two did not get disease). Disease due to eating raw meat is not analyzed under the objectives of this review. |

| **Quality appraisal** | **Authors’ judgment** | **Support for judgment** |
| --- | --- | --- |
| 1. Were patient characteristics adequately reported? | Yes | Patient’s characteristics described |
| 2. Was there some effort to trace all contacts from the index case? | Yes | Detailed |
| 3. Were the methods used for tracing contacts adequate? | Yes | Detailed description of contact tracing given |
| 4. Were the laboratory methods used for defining a confirmed case of plague reliable? | Yes | Indirect hemagglutination |
| 5. Was the route of transmission plausible? | Yes | Direct contact with breaks of skin described |
| 6. Was the cause-effect of transmission plausible? | Yes | With presence of axillary lymphadenitis on the same side of the cut with direct handling |

| **Sagiev 2019** | |
| --- | --- |
| **Setting** | **Country:** Kazakhstan  **Study dates:** 1974-2003 |
| **Source of infection** | **Source of infection:**  A) slaughter of camel  B) cutting carcasses of hare  C) contact with corpse of hare  D) cutting a sick saiga  **Plague diagnosis:**  D) plague microbes isolated from body of saiga  **Other possible sources of infection:** not reported |
| **Exposure to the source of infection** | **Description of exposure:**  C) fed the eagle with corpse of hare  **Duration of exposure:** not reported  **Time between death of the human/animal and contact with the infected human:** not reported |
| **Persons infected** | **Number:** 12 in total: A) 8; B) 2; C) 1; D) 1  **Age, sex:** C) 13 years; others not reported  **Profession:** not reported  **Signs and symptoms:** not reported  **Form of plague:** not reported  **Plague diagnosis:** D) plague microbes isolated  **Outcome:** not reported |
| **Route of transmission** | **Attributed route of transmission by the study author:** not reported  **Plausibility of the cause-effect transmission as per the study authors:** not applicable |
| **Other relevant notes** | **Cases exposed to the same source but not infected:** none reported |

| **Quality appraisal** | **Authors’ judgment** | **Support for judgment** |
| --- | --- | --- |
| 1. Were patient characteristics adequately reported? | No | Patient characteristics not described |
| 2. Was there some effort to trace all contacts from the index case? | Unknown | Not reported |
| 3. Were the methods used for tracing contacts adequate? | Not applicable | Not applicable |
| 4. Were the laboratory methods used for defining a confirmed case of plague reliable? | Unknown | Only one case has details of laboratory diagnosis |
| 5. Was the route of transmission plausible? | Unknown | While direct handling including cutting cadavers is a possibility, there are no details provided on route of transmission, and on form of plague. |
| 6. Was the cause-effect of transmission plausible? | Unknown | Absence of details on type of plague is a limitation |

| **Von Reyn 1976** | |
| --- | --- |
| **Setting** | **Country:** New Mexico  **Study dates:** February 1974 |
| **Source of infection** | **Source of infection:** coyote  **Plague diagnosis:** fluorescent antibody test positive from spleen and bone marrow of carcass  **Other possible sources of infection:** patient cannot recall insect bites |
| **Exposure to the source of infection** | **Description of exposure:** skinned the animal and carried the pelt. Had forearm laceration and nailbed exposed during skinning. Authors raise the possibility of animal not being dead at the time of exposure and being weak.  **Duration of exposure:** not reported  **Time between death of the human/animal and contact with the infected human:** not reported |
| **Persons infected** | **Number:** 1  **Age, sex:** 11 years, male  **Profession:** not reported  **Signs and symptoms:** fever, right axillary lymphadenopathy and wound on right middle digit. Later fever, neck stiffness and lethargy.  **Form of plague:** bubonic plague, plague meningitis  **Plague diagnosis:** lymph nodes and cerebrospinal fluid had positive culture for plague bacilli  **Outcome:** recovery |
| **Route of transmission** | **Attributed route of transmission by the study author:** direct contact with break in skin  **Plausibility of the cause-effect transmission as per the study authors:** only the person with breach in the skin developed the disease |
| **Other relevant notes** | **Cases exposed to the same source but not infected:** patient’s friend who joined skinning the cadaver. Several members at two households handled the skin.  Passive plague hemagglutination negative in ten exposed people |

| **Quality appraisal** | **Authors’ judgment** | **Support for judgment** |
| --- | --- | --- |
| 1. Were patient characteristics adequately reported? | Yes | All patient characteristics described in detail |
| 2. Was there some effort to trace all contacts from the index case? | Yes | Contacts who were not infected were described in detail including assessment of serology status |
| 3. Were the methods used for tracing contacts adequate? | Yes | Contacts who helped to skin the coyote were traced |
| 4. Were the laboratory methods used for defining a confirmed case of plague reliable? | Yes | Diagnosed using isolation of the organism |
| 5. Was the route of transmission plausible? | Yes | Direct contact with breaks of skin described |
| 6. Was the cause-effect of transmission plausible? | Yes | Axillary buboes were noted on the same side of skin break |

| **Wong 2009** | |
| --- | --- |
| **Setting** | **Country:** the United States  **Study dates:** November 2007 |
| **Source of infection** | **Source of infection:** Mountain Lion  **Plague diagnosis:** samples from the liver and submandibular lymph node: PCR positive for *Y pestis*. Subcapsular sinuses of a submandibular lymph node: abundant gram negative bacilli and *Y pestis* intensely identified by immunohistochemistry stain*.* Liver and brain samples: plague bacilli detected by immunohistochemistry  **Other possible sources of infection:** a search did not identify any other sources |
| **Exposure to the source of infection** | **Description of exposure:**  “The biologist carried the carcass for approximately 1 km to his vehicle and then into his garage, where he performed a necropsy with his bare hands; there is no evidence he wore a mask or other personal protective equipment.”  The necropsy of the animal included opening the animal’s thoracic cavity, found filled with blood, and transecting the vertebral column.  Archived specimens of the animal by the biologist included hide, 2 paws, skinned head and liver.  **Duration of exposure:** Time of transport of the carcass for 1km, then in his car.  Then estimated 2.5h of examination.  **Time between death of the human/animal and contact with the infected human:** the infected case first evidenced-contact (photography) is 35 hours after the death of the mountain lion. Death of the animal was identified from a ‘mortality signal’ defined as no movement for 6 hours, transmitted from the animal’s radio-collar. |
| **Persons infected** | **Number:** 1  **Age, sex:** 37 years, male  **Profession:** wildlife biologist  **Signs and symptoms:** fever, chills, nausea, myalgias, cough, blood-tinged sputum few hours after exposure  **Form of plague:** primary pneumonic plague  **Plague diagnosis:** “intravascular *Y. pestis* antigens [identified] by immunohistochemistry in multiple tissue samples, including samples of the lung, liver, heart, pharynx, and brain”  - Confluent plague bacilli admixed with an acute inflammatory infiltrate in the lung; inflammation absent from other infected organs.  - “Culture of patient tissue samples (lung and liver) yielded *Y. pestis,* as confirmed by bacteriophage-lysis testing.”  **Outcome:** Death |
| **Route of transmission** | **Attributed route of transmission by the study author:** inhalation of aerosols generated while handling the infected animal.  **Plausibility of the cause-effect transmission as per the study authors:**  “The presence of heavy intra-alveolar inflammation admixed with confluent plague bacilli—in conjunction with the complete absence of inflammation in other infected organs—provides strong evidence that the lungs were the primary site of infection and that septicemia occurred secondarily”  “Other findings consistent with an aerosol exposure include the development of cough and blood-tinged sputum within hours of symptom onset, consolidation of the right lung, and the absence of buboes on clinical and postmortem examination.”  “Isolates of *Y. pestis* cultured from the mountain lion’s tissues were subtyped by pulsed-field gel electrophoresis (PFGE) and found to be indistinguishable from isolates recovered from the biologist”, which supports the mountain lion as the source of the biologist’s infection. |
| **Other relevant notes** | **Cases exposed to the same source but not infected:** none |

| **Quality appraisal** | **Authors’ judgment** | **Support for judgment** |
| --- | --- | --- |
| 1. Were patient characteristics adequately reported? | Yes | All patient characteristics described in detail |
| 2. Was there some effort to trace all contacts from the index case? | Yes | Through interviews, photographs, and cellular phone records |
| 3. Were the methods used for tracing contacts adequate? | Yes | Refer above |
| 4. Were the laboratory methods used for defining a confirmed case of plague reliable? | Yes | Multiple methods used |
| 5. Was the route of transmission plausible? | Yes | Through aerosols |
| 6. Was the cause-effect of transmission plausible? | Yes | Clinical picture strongly suggests the route of exposure |

| **Wu 2009** | |
| --- | --- |
| **Setting** | **Country:** China  **Study dates:** 1975 to 2007 |
| **Source of infection** | **Source of infection:** Tibetan sheep  **Plague diagnosis:** isolated *Yersinia pestis* strains  **Other possible sources of infection:** not reported |
| **Exposure to the source of infection** | **Description of exposure:** humans infected by flaying, eating or touching the cadaver of Tibetan sheep  **Duration of exposure:** not reported  **Time between death of the human/animal and contact with the infected human:** not reported |
| **Persons infected** | **Number:** 25  **Age, sex:** 9 cases <20 years, 8 cases between 20 and 45 years, 8 cases >45 years  11 males; 14 females.  **Profession:** not reported  **Signs and symptoms:** not reported  **Form of plague:** 12 primary bubonic, 6 primary pneumonic, 3 primary septicaemic, 4 primary intestinal  **Plague diagnosis:** by trade standard (WS279-2008):  a. sudden high fever to be determined: symptoms + visited plague foci in past 10 days.  b. probable case: clinical symptoms + contact history + F1 antigen positive by TDR, IHA or ELISA.  c. laboratory confirm case:  a. or b.  + isolated *Y. pestis* strains      a. + 4 folds increased of F1 antibody  **Outcome:** recovery for 12 cases, 13 deaths |
| **Route of transmission** | **Attributed route of transmission by the study author:** not reported  **Plausibility of the cause-effect transmission as per the study authors:** not applicable |
| **Other relevant notes** | **Cases exposed to the same source but not infected:** none reported |

| **Quality appraisal** | **Authors’ judgment** | **Support for judgment** |
| --- | --- | --- |
| 1. Were patient characteristics adequately reported? | Yes | Basic information reported |
| 2. Was there some effort to trace all contacts from the index case? | Unknown | Not reported |
| 3. Were the methods used for tracing contacts adequate? | Not applicable | Not applicable |
| 4. Were the laboratory methods used for defining a confirmed case of plague reliable? | Yes | Diagnosed using isolation of the organism |
| 5. Was the route of transmission plausible? | Yes | Cases of bubonic and pneumonic plague can be the resulted from close direct contact and inhalation of infected aerosols from the infected cadaver. Cases of intestinal plague are likely to be the result of eating contaminated meat. |
| 6. Was the cause-effect of transmission plausible? | Partial | There is not enough information given to exclude that some cases were due to human to human transmission. |

| **Zhang 2007** | |
| --- | --- |
| **Setting** | **Country:** China  **Study dates:** 1958 to 2005 |
| **Source of infection** | **Source of infection:** animals or human with plague; the original infection sources were live animals or cadaver which were infected with plague  **Plague diagnosis:** not reported  **Other possible sources of infection:** not reported |
| **Exposure to the source of infection** | **Description of exposure:**  24 patients were infected by flaying, eating or touching a marmot or cat.  32 patients were infected by contacting with plague patients or human cadaver (not mentioned respective numbers).  **Duration of exposure:** not reported  **Time between death of the human/animal and contact with the infected human:** not reported |
| **Persons infected** | **Number:** 56 (Out of 64 in which 8 cases with unknown route of transmission were included)  **Age, sex:** age ranged between 1 and 69 years; 49 males, 15 females.  **Profession:** not reported  **Signs and symptoms:** not reported  **Form of plague:** 45% pneumonic plague, 33% bubonic plague, 8% septicaemic plague  **Plague diagnosis:**  Laboratory confirm cases:  a. isolated the strain (26 patients)  b. clinical symptoms + F1 antibody titer ≥ 1:20 (by IHA, indirect hemagglutination assay) (2 patients)  c. clinical symptoms + F1 antigen positive by RIHA (reverse indirect hemagglutination assay) (1patient)  Clinical cases: symptoms + epidemiological evidence (35 patients)  **Outcome:** not reported |
| **Route of transmission** | **Attributed route of transmission by the study author:** not reported  **Plausibility of the cause-effect transmission as per the study authors:** not applicable |
| **Other relevant notes** | **Cases exposed to the same source but not infected:** not reported |

| **Quality appraisal** | **Authors’ judgment** | **Support for judgment** |
| --- | --- | --- |
| 1. Were patient characteristics adequately reported? | Partial | Inadequate information on some aspects |
| 2. Was there some effort to trace all contacts from the index case? | Unknown | Not reported |
| 3. Were the methods used for tracing contacts adequate? | Not applicable | Not applicable |
| 4. Were the laboratory methods used for defining a confirmed case of plague reliable? | Partial | Diagnosis was based on clinical and epidemiological findings for 35 cases |
| 5. Was the route of transmission plausible? | Unknown | Very limited information to make this judgment. While it is plausible, no data reported on the route between the contact with infected persons/animals/cadavers and the different forms of plague. |
| 6. Was the cause-effect of transmission plausible? | Partial | There is not enough information given to exclude that some cases were due to human to human transmission. |
